# Risk factors for SARS-CoV-2 seroprevalence following the first pandemic wave in UK healthcare workers in a large NHS Foundation Trust

**DOI:** 10.1101/2021.07.07.21260151

**Authors:** David Hodgson, Hayley Colton, Hailey Hornsby, Rebecca Brown, Joanne Mckenzie, Kirsty L. Bradley, Cameron James, Benjamin B. Lindsey, Sarah Birch, Louise Marsh, Steven Wood, Martin Bayley, Gary Dickson, David C. James, Martin J.H. Nicklin, Jon R. Sayers, Domen Zafred, Sarah L. Rowland-Jones, Goura Kudesia, Adam Kucharski, CMMID COVID-19 Working Group, Thomas C. Darton, Thushan I. de Silva, Paul J. Collini

## Abstract

**Background:** We aimed to measure SARS-CoV-2 seroprevalence in a cohort of healthcare workers (HCWs) during the first UK wave of the COVID-19 pandemic, explore risk factors associated with infection, and investigate the impact of antibody titres on assay sensitivity.

**Methods:** HCWs at Sheffield Teaching Hospitals NHS Foundation Trust (STH) were prospectively enrolled and sampled at two time points. SARS-CoV-2 antibodies were tested using an in-house assay for IgG and IgA reactivity against Spike and Nucleoprotein (sensitivity 99·47%, specificity 99·56%). Data were analysed using three statistical models: a seroprevalence model, an antibody kinetics model, and a heterogeneous sensitivity model.

**Findings:** As of 12th June 2020, 24·4% (n=311/1275) HCWs were seropositive. Of these, 39·2% (n=122/311) were asymptomatic. The highest adjusted seroprevalence was measured in HCWs on the Acute Medical Unit (41·1%, 95% CrI 30·0–52·9) and in Physiotherapists and Occupational Therapists (39·2%, 95% CrI 24·4–56·5). Older age groups showed overall higher median antibody titres. Further modelling suggests that, for a serological assay with an overall sensitivity of 80%, antibody titres may be markedly affected by differences in age, with sensitivity estimates of 89% in those over 60 years but 61% in those ≤30 years.

**Interpretation:** HCWs in acute medical units working closely with COVID-19 patients were at highest risk of infection, though whether these are infections acquired from patients or other staff is unknown. Current serological assays may underestimate seroprevalence in younger age groups if validated using sera from older and/or more symptomatic individuals.

**Research in context:** *Evidence before this study:* We searched PubMed for studies published up to March 6th 2021, using the terms “COVID”, “SARS-CoV-2”, “seroprevalence”, and “healthcare workers”, and in addition for articles of antibody titres in different age groups against coronaviruses using “coronavirus”, “SARS-CoV-2, “antibody”, “antibody tires”, “COVID” and “age”. We included studies that used serology to estimate prevalence in healthcare workers. SARS-CoV-2 seroprevalence has been shown to be greater in healthcare workers working on acute medical units or within domestic services. Antibody levels against seasonal coronaviruses, SARS-CoV and SARS-CoV-2 were found to be higher in older adults, and patients who were hospitalised.

*Added value of this study:* In this healthcare worker seroprevalence modelling study at a large NHS foundation trust, we confirm that those working on acute medical units, COVID-19 “Red Zones” and within domestic services are most likely to be seropositive. Furthermore, we show that physiotherapists and occupational therapists have an increased risk of COVID-19 infection. We also confirm that antibody titres are greater in older individuals, even in the context of non-hospitalised cases. Importantly, we demonstrate that this can result in age-specific sensitivity in serological assays, where lower antibody titres in younger individuals results in lower assay sensitivity.

*Implications of all the available evidence:* There are distinct occupational roles and locations in hospitals where the risk of COVID-19 infection to healthcare workers is greatest, and this knowledge should be used to prioritise infection prevention control and other measures to protect healthcare workers. Serological assays may have different sensitivity profiles across different age groups, especially if assay validation was undertaken using samples from older and/or hospitalised patients, who tend to have higher antibody titres. Future seroprevalence studies should consider adjusting for age-specific assay sensitivities to estimate true seroprevalence rates.

**Author Contributions:** 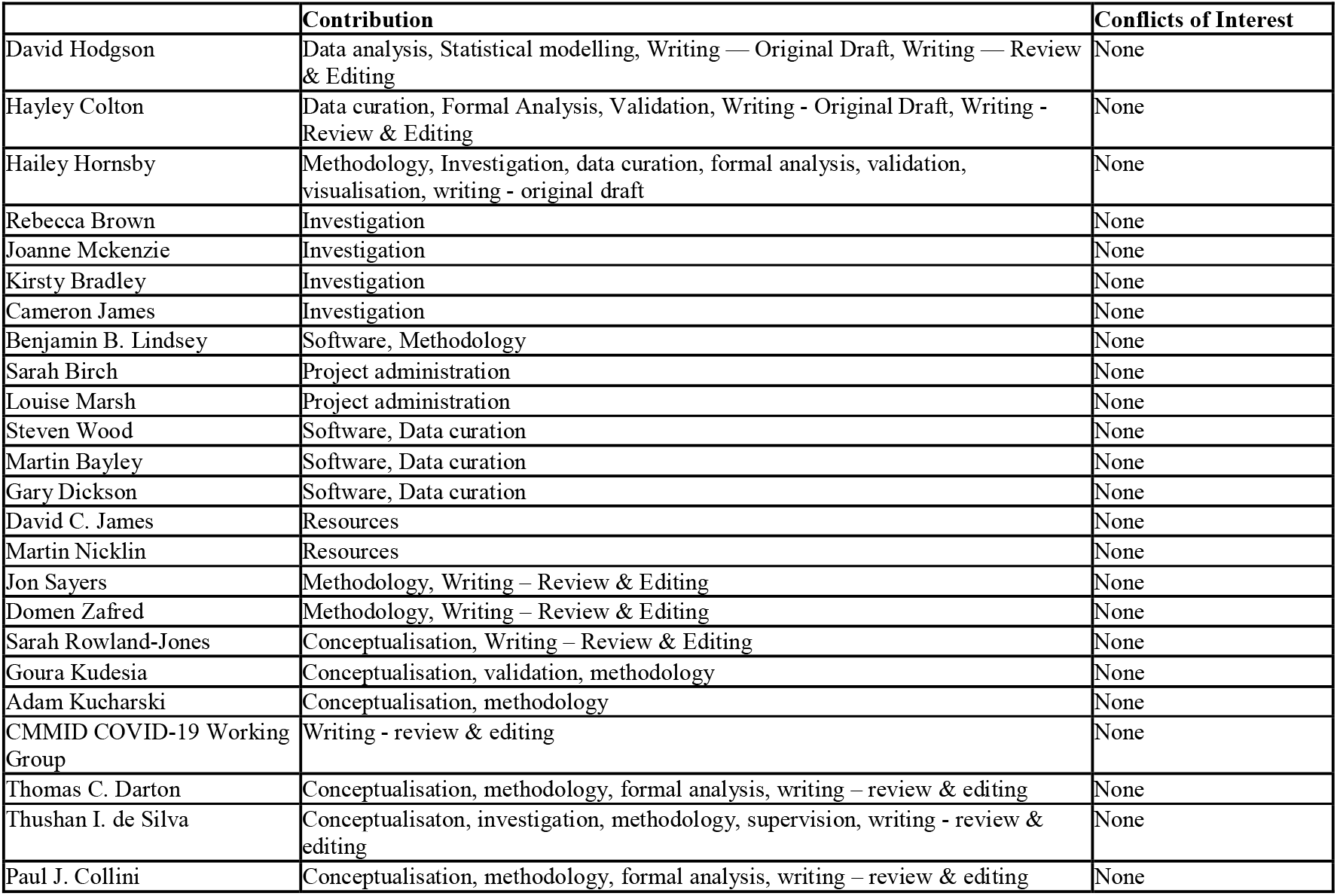

## Introduction

Throughout the SARS-CoV-2 pandemic, healthcare workers (HCWs) have been at increased risk of acquiring COVID-19.^1,2^ The true number of HCWs exposed to SARS-CoV-2 to date is not fully established, particularly during the first wave of the pandemic in the UK. At first, methods for estimating this number included extrapolating from sickness reporting or work absenteeism, although these were unlikely to be reliable for multiple reasons including heightened concern of infecting workplace colleagues or patients.^3^ Confirmation by molecular testing provided a more accurate picture of confirmed cases when it was available, although access to nucleic acid amplification testing (NAAT) was restricted early on in the UK pandemic to hospitalised patients once community testing ceased on 12 March 2020.^4^

An alternative population-level approach is to look at the number who have detectable antibodies against SARS-CoV-2 antigens at one or more timepoints. Such HCW seroprevalence studies may offer a more comprehensive measure of the true numbers infected over time and are less affected by symptom-activated testing pathways.^5–8^ These studies may be useful for characterising the risk factors for SARS-CoV-2 exposure in healthcare settings.

The accuracy of seroprevalence measurement depends on the characteristics of antibody evolution and thus sampling time relative to infection onset, immunoglobulin isotype, antigenic target and assay performance.^9–14^ The performance of serological assays has mostly been evaluated using samples from hospitalised patients, leaving it unclear how they perform with the lower antibody levels likely seen with milder and asymptomatic COVID-19 disease.^10,12^ While studies on antibody responses to various coronaviruses suggest antibody levels are greater in older people, it is unclear whether this is due to higher exposure risk or augmented humoral responses due to factors such as heterologous boosting from prior infections.^15–19^ Also not explored is whether the differences in antibody titres across ages may result in age-specific differences in antibody assay sensitivity, which may be a significant confounder in population seroprevalence studies.

In this study we aimed to measure the proportion of healthcare workers at Sheffield Teaching Hospitals NHS Foundation Trust (STH), UK, who were infected with SARS-CoV-2 during the first wave of the pandemic (from March 2020 to June 2020) by estimating the seroprevalence of SARS CoV-2 in a large cohort of HCW. We used statistical models to explore risk factors associated with infection in non-hospitalised HCW, as well as antibody kinetics and the potential impact of varying antibody titres across age groups on assay sensitivity.

## METHODS

### Background and setting

STH offers secondary and tertiary hospital care across four sites in South Yorkshire, UK. STH has 1,669 inpatient beds and employs a total of around 18,500 staff, serving a population of 640,000.^20^ The first patient at STH with confirmed COVID-19 was admitted on 23 February 2020. From 17 March 2020, symptomatic staff testing using self-collected combined nose and throat swabs for SARS-CoV-2 NAAT was initiated, and on the same day Public Health England (PHE) de-escalated personal protective equipment (PPE) recommendations for HCWs caring for inpatients with suspected or confirmed COVID-19 (i.e. from ‘Level 3 Airborne’ to ‘Level 2 Droplet’ for routine care, see Supplementary Information for PPE level definitions). Universal ‘Level 2 Droplet’ PPE for all inpatient and outpatient care began on 08 April 2020. Asymptomatic screening of staff working in outbreak and high-risk areas was rolled out from 18 May 2020 using self-collected combined nose and throat swabs for SARS-CoV-2 NAAT, and STH policy was changed on 15 June 2020 to mandate all staff to wear surgical face masks whilst on hospital grounds.

### Recruitment & Consent

Between the 13th and 18th May 2020, all STH staff on the communications email distribution list (n=17,757) were contacted to invite them to take part in the COVID-19 <u>H</u>umoral Immun<u>E</u> <u>R</u>esp<u>O</u>nses in front-line health care workers study (HERO), emphasising a focus on recruiting clinical and non-clinical staff working in inpatient areas, in addition to an alert on the staff intranet page. To aid recruitment in areas with limited computer access (e.g. domestic services staff), posters were displayed in work areas and three sessions were held to help enrol staff who had expressed an interest in the study. Immediately following electronic informed consent, participants received a study email with a link to an online questionnaire which used a unique study number and retrospectively collected self-reported data on age, gender, ethnicity, job role, and working environment since start of pandemic (to stratify risk of direct contact with COVID-19 patients; including “COVID-19 zone” - defined in Supplementary Information). The history and timing of any possible or confirmed COVID-19 illness since the beginning of February 2020 were collected into the following three groups: diagnosed COVID-19 and NAAT confirmed, clinically diagnosed COVID-19 but NAAT not performed, and self-reported symptomatic (Supplementary Information). We defined these three groups as “symptomatic”, as asymptomatic testing was only introduced after the recruitment period in our study. Those reporting no diagnosis or symptoms consistent with COVID-19 from February 2020 to date of recruitment were defined as “asymptomatic”. Participants were invited to attend two visits, four weeks apart, for serum sampling. All participants who agreed to be contacted were sent their first and second serology result by email following their second visit, with accompanying information about interpreting antibody test results.

### SARS-CoV-2 serology

Serum samples were tested for IgG and IgA reactivity to two SARS-CoV-2 proteins - full-length extracellular domain (amino acids 14-1213) of Spike glycoprotein, including a replacement of the furin cleavage site R684-R689 by a single alanine residue and replacement of K986-V987 by PP, produced in mammalian cells, and full-length untagged Nucleoprotein (NCP) produced in *E. coli* (Uniprot ID P0DTC9 (NCAP_SARS2), https://www.uniprot.org/uniprot/P0DTC9).21–^23^ High binding microtitre plates (Immulon 4HBX; Thermo Scientific, 6405 or Nunc MaxiSorp; Thermo Scientific, 442404) were coated overnight with either Spike or NCP protein diluted in phosphate buffered saline (PBS), washed with 0·05% PBS-Tween, and blocked for 1 hour with 200 µL per well casein blocking buffer. Prior to testing, serum samples were heat inactivated at 56L for 30 minutes. Following assay optimisation (Supplementary Information), samples were diluted 1:200 for the IgG assay or 1:100 for the IgA assay. Plates were emptied and 100 µL per well of samples and controls loaded and incubated for 2 hours. Plates were washed and loaded with 100 µL per well of goat anti-human IgG-HRP conjugate (Thermo Scientific, A6907) at 1:500 for the IgG assay, or goat anti-human IgA-HRP conjugate (Invitrogen, 11594230) at 1:1000 for the IgA assay, for one hour. Plates were washed and developed for 10 minutes with 100 µL per well TMB substrate (KPL, 5120-0074). Development was stopped with 100 µL per well HCl Stop solution (KPL, 5150-0021), and absorbance read at 450nm. All steps were performed at room temperature.

A calibration curve of sera pooled from two convalescent SARS-CoV-2 NAAT confirmed patients with high antibody titres for both spike and NCP was included on plates to allow quantification of antibody concentrations. The curve was generated by serially diluting in 1·75× steps from a starting concentration of 1:200 for the IgG assay or 1:100 for the serum IgA assay (to match sample dilution), to create a 12-well dilution series. Initially, an arbitrary value of 1000 antibody units was assigned to the most concentrated well of the series, the curve was then used to extrapolate antibody unit values for the samples on the plate. For the IgG assay, this calibration curve was later run in parallel with the WHO International Standard for anti-SARS-CoV-2 immunoglobulin when this became available (NIBSC, 20/136), to allow conversion of our antibody units to the WHO-recommended universal antibody units (Supplementary Information). Thus for the IgG assay, antibody values are given in WHO antibody units, while for the IgA assay they are given as our arbitrary antibody units.

### Sample Size

To meet the primary objective of the study, measurement of the overall seroprevalence of SARS-CoV-2 IgG, we calculated a sample size of 1,000 STH staff would provide a +/-1·4% precision based on a seroprevalence estimate that ∼4% of the UK population may have been infected by April 2020, with a two-sided 95% confidence interval (with n=753, Binomial exact 95%CI has been estimated to be 2·7-5·6%).^24^

### Statistical modelling overview

We considered three statistical models, i) a seroprevalence model, ii) an antibody kinetics model, and iii) a heterogeneous sensitivity model. For the seroprevalence model, we used the serostatus of all samples at their first blood draw in a sensitivity and specificity adjusted Bayesian multilevel logistic regression model. Using seropositivity as the binary response variable, we considered three different model subtypes with varying primary exposures; job location, contact with COVID-19 patients, and job type (Supplementary Information). All three of these models were evaluated either unadjusted (primary exposure only) or adjusted (primary exposure with age, gender, and ethnicity). Age group categories considered were <30, 30-39, 40-49, 50-59, and 60 years plus; gender categories considered included Female and Male, and Ethnicity categories included White, Black, Asian and Other (which includes “Prefer not to say”). After fitting, we performed a post-stratification analysis to estimate the seroprevalence of each covariate by averaging out the contribution of the other covariates in the model. In addition to the seroprevalence model, we fitted a symptomatic prevalence model, where the data used were seropositive persons only, and the binary response variable was asymptomatic or symptomatic infection (Supplementary Information).

For the antibody kinetics model, we included samples from persons who were seropositive at both their first and second bleed in a Bayesian multilevel linear regression model. There are two parts of the model; the first used antibody log2 antibody units (logAU) at the first blood draw as the response variable and the second used the change in antibody titre at the follow up bleed (median 28 days) as the response variable. Both models used age, ethnicity, gender and symptom severity (asymptomatic or symptomatic) as covariates and each model was run separately for four different antibody-antigen combinations; Spike-IgG, NCP-IgG, Spike-IgA, NCP-IgA. The time until seroreversion was calculated for each covariate group and antibody-antigen interactions by i) sampling a starting titre value and a rate of decline from the two models, and then ii) calculating the time until the minimum observed antibody value was reached for that antibody-antigen interaction, assuming a continuous rate of decrease.

In our heterogeneous sensitivity and specificity model, we were interested in exploring how estimates for sensitivity and specificity taken from our assay validation dataset generalise to covariate groups (e.g. age) within our dataset. The samples used for assay validation consisted of NAAT-confirmed SARS-CoV-2 samples from the current serosurvey, in addition to serum from NAAT-confirmed hospitalised cases at STH. To model the generalisability of these performance measures, we compared the seropositivity classification of our study dataset using our in-house ELISA, with the predicted seropositivity classification from hypothetical serological assays with a quoted sensitivity and specificity. Our model considers the different distribution of the *A*_450_ values in the assay validation and HERO study datasets to model how reliably quoted performance measures generalise. Using the assay validation dataset, we estimated the *A*_450_ cut-off value for a range of chosen sensitivity values, and then used this *A*_450_ cut-off to classify seropositivity in the study dataset. We then estimated the implied sensitivity on the HERO dataset by comparing seropositivity classification based on the estimated *A*_450_ cut-off value, with the seropositivity classification from our in-house ELISA (which for ease of comparison, we assume represents the maximum possible sensitivity and specificity (i.e. 100%) in this model). This framework allowed us to estimate the hypothetical performance of serological assays reported in the literature, on our HCW dataset. As any covariate (e.g. age) which displays variability in antibody distribution will also have variation in their *A*_450_ distribution, we estimated implied covariate-specific sensitivity on the study dataset.

All Bayesian regression analysis was performed in R version 4.0.2 and cmdstanr version 0.2.0. Full details of the model outline and implementation are given in Supplementary Information. An R package containing all the analysis in this study is available at https://github.com/dchodge/hero-study.

### Regulatory review

Following internal scientific review, local R&D (5 May 2020 ref: STH21394) and HRA and Health and Care Research Wales (HCRW) approval were given (29 April 2020 ref: 20/HRA/2180, IRAS ID: 283461). Anonymised serum samples from hospitalised COVID-19 patients and serum collected prior to 2017 during routine clinical care were used for assay validation according to guidance and approval from STH R&D office.

## RESULTS

### Serology assay development

The assay was validated using serum samples collected from 190 SARS-CoV-2 NAAT-confirmed cases, and 675 patients sampled prior to 2017. SARS-CoV-2 NAAT-confirmed validation samples used were from hospitalised patients (n=52) and healthcare workers with mild infections recruited in this study (n=138). Sample time points were between 14 to 120 days after NAAT-positive results. Pre-pandemic samples were selected from patients with other confirmed respiratory viruses, acute cytomegalovirus (CMV) or Epstein Barr (EBV) infections, HIV-1 infection, and patients undergoing haemodialysis, in order to assess cross-reactivity or non-specific reactivity (Table S1).

Thresholds for defining a reactive sample in spike (*A*_450_ 0·175) and NCP (*A*_450_ 0·1905) were set to optimise the sensitivity of each assay. Given the IDSA guidance for ensuring a specificity of at least 99·5% in assays used for SARS-CoV-2 seroprevalence studies, specificity was enhanced by defining a SARS-CoV-2 seropositive sample as one where both spike and NCP were reactive.^25^ This resulted in a sensitivity of 99·47% (95% confidence interval (CI) 97·10% - 99·99%) and specificity of 99·56% (95% CI 98·71% - 99·91%) for our in house IgG ELISA (Fig S1).. Waning of IgA responses more rapidly following SARS-CoV-2 infection meant defining positive and negative samples based on the convalescent set of sera we used for assay validation was challenging. We also observed more background reactivity for IgA compared to IgG in pre-pandemic samples. We therefore opted to use our optimised spike and NCP IgA ELISA purely for comparing IgA levels in individuals classified as seropositive in our IgG assay (Fig S2). Antibody units at each given dilution of the calibration curve are shown in Table S2.

### Registration and Study Visits

Of those contacted, 1478 STH staff registered to take part between 13th May and 5th June 2020 (Fig. S3). Of these, 1277 attended for a first visit and blood draw (V1) between 15^th^ May 2020 and 12^th^ June 2020. As 2 samples were contaminated in transit, we obtained a valid serostatus for 1275 samples. The second visits and blood draw were conducted between 15^th^ June and 10^th^ July 2020, and 1174 attended for a second visit and blood draw (V2) (Fig. S3 and S4).

### Demographics, Job Role, Work Locations, and Environment

Of the 1275 participants included in the analysis for V1, the majority were female (n=1008/1275, 79·1%) and most participants described their ethnicity as white (n=1130/1275, 88·6%), with no other ethnic group representing more than 3% (Table 1). Nurses (433/1275, 33·9%), doctors (232/1275, 18·2%), health care assistants (163/1275, 12·8%), and domestic services staff (136/1275, 10·7%) made up the largest proportion of job roles represented in the study. Almost half (593/1275, 46·5%) of HCWs participating in our study worked in parts of the hospital managing acute COVID-19 admissions including the Emergency Department (ED), Acute Medical Unit (AMU), Critical Care, and inpatient medical wards (respiratory, geriatric care, infectious diseases). Most of our participants reported that they had been working in areas with the highest degree of COVID-19 patient contact (red zones), either most days (n=423/1275, 33·2%) or occasionally (n=305/1275, 23·9%).

**Table 1.** Characteristics and serostatus of recruited participants who had a valid baseline result.

|  | Recruited with an initial valid serostatus | Seropositive at V1 (%) | Asymptomatic (% of seropositive at V1) | Completed both V1 and V2 (% of recruited) | Seroincident cases (% of seronegative at V1) |
| --- | --- | --- | --- | --- | --- |
| Total | 1275 | 311 (24.4) | 122 (39.2) | 1166 (87.5) | 16 (1.7%) |
| **Job location** |  |  |  |  |  |
| Emergency Department (ED) | 103 | 26 (25.2) | 13 (50.0) | 90 (87.4) | 1 (1.2) |
| Acute Medical Unit (AMU) | 83 | 38 (45.8) | 17 (44.4) | 66 (79.5) | 0 (0.0) |
| Critical Care | 100 | 18 (18.0) | 7 (38.9) | 95 (95.0) | 0 (0.0) |
| Geriatric Care | 23 | 3 (13.0) | 1 (33.3) | 22 (95.7) | 1 (5.0) |
| Infectious Disease Ward | 139 | 26 (18.7) | 11 (42.3) | 121 (87.1) | 7 (6.2) |
| Other | 664 | 157 (23.6) | 56 (35.7) | 621 (93.5) | 2 (0.3) |
| Respiratory Geriatric Ward | 92 | 27 (29.3) | 10 (37.0) | 85 (92.4) | 2 (3.1) |
| Respiratory Ward | 58 | 13 (22.4) | 5 (38.5) | 54 (93.1) | 2 (4.4) |
| **Job role** |  |  |  |  |  |
| Admin | 127 | 26 (20.5) | 12 (46.2) | 118 (92.9) | 1 (0.9) |
| Allied medical<sup>1</sup> | 38 | 0 (0.0) | — | 37 (97.4) | 0 (0.0) |
| Domestic services | 136 | 39 (28.7) | 24 (61.5) | 127 (93.4) | 4 (4.1) |
| Healthcare assistants | 163 | 39 (23.9) | 21 (53.8) | 140 (85.9) | 3 (2.4) |
| Doctors | 232 | 52 (22.4) | 18 (34.6) | 211 (90.9) | 0 (0.0) |
| Nurses | 433 | 116 (26.7) | 34 (29.3) | 391 (90.3) | 7 (2.2) |
| Other | 31 | 5 (16.1) | 2 (40.0) | 29 (93.5) | 0 (0.0) |
| Pharmacists | 35 | 8 (22.8) | 5 (62.5) | 33 (94.3) | 1 (3.7) |
| Occupational and physiotherapists | 33 | 15 (45.5) | 3 (20.0) | 33 (100.0) | 0 (0.0) |
| Radiographers | 42 | 9 (21.4) | 2 (22.2) | 42 (100.0) | 0 (0.0) |
| **COVID-19 zone<sup>2</sup>** |  |  |  |  |  |
| 1 (lowest COVID-19 contact) | 104 | 22 (21.2) | 10 (54.5) | 96 (92.3) | 1 (1.2) |
| 2 | 248 | 50 (20.2) | 27 (46.0) | 232 (93.5) | 0 (0.0) |
| 3 | 41 | 7 (17.1) | 6 (14.3) | 39 (95.1) | 1 (2.9) |
| 4 | 153 | 35 (22.9) | 24 (31.4) | 142 (92.8) | 1 (0.8) |
| 5 | 305 | 69 (22.6) | 46 (33.3) | 280 (91.8) | 3 (1.3) |
| 6 (highest COVID-19 contact) | 423 | 128 (30.3) | 76 (40.6) | 376 (88.9) | 10 (3.4) |
| **Age (years)** |  |  |  |  |  |
| <30 | 267 | 67 (25.1) | 32 (47.8) | 236 (88.4) | 6 (3.0) |
| 30–39 | 306 | 69 (22.5) | 22 (31.8) | 279 (91.2) | 5 (2.1) |
| 40–49 | 314 | 72 (22.9) | 29 (40.2) | 293 (93.3) | 2 (0.8) |
| 50–59 | 314 | 76 (24·2) | 28 (36·8) | 288 (91·7) | 1 (0·4) |
| 60+ | 74 | 27 (36·5) | 11 (40·7) | 70 (94·6) | 2 (4·3) |
| **Ethnicity** |  |  |  |  |  |
| White | 1130 | 281 (24·9) | 108 (38·4) | 1035 (91·6) | 15 (1·8) |
| Black/Black British | 33 | 6 (18·2) | 3 (50·0) | 30 (90·9) | 0 (0·0) |
| Asian/Asian British | 76 | 17 (22·4) | 7 (41·2) | 70 (92·1) | 1 (1·7) |
| Other | 33 | 7 (21·2) | 4 (57·1) | 30 (90·9) | 0 (0·0) |
| **Gender<sup>3</sup>** |  |  |  |  |  |
| Female | 1008 | 253 (24·1) | 105 (41·5) | 922 (91·5) | 14 (1·9) |
| Male | 265 | 58 (21·9) | 17 (29·3) | 242 (91·3) | 2 (0·9) |

1. Allied Medical includes Speech and Language Therapists, Cardiac Physiologists, Dental Hygienists, Dietitians, ECG technicians, Orthotists, Podiatrists, Rehabilitation assistants 2. COVID-19 Zones are defined in supplementary information 3. Participants were able to define their gender as non-binary, transgender or could choose not to disclose

### Unadjusted Seroprevalence

Analysis of samples collected at the initial visit revealed that 24·4% (n=311/1275) of HCWs participating in our study were seropositive by 12th June 2020 (Table 1). Of these, 39·2% (n=122/311) did not report an illness consistent with COVID-19 prior to serological testing. The second blood draw occurred a median of 28 days following the first visit (IQR 27-31, range 12 – 52 days) and 1166 had a valid V2 serology result (Fig. S3). Comparison of serology data with the initial blood draw demonstrated that 16 out of 964 participants had seroconverted (i.e. reactive against both spike (*A*_450_ ≥0·1905) and NCP (*A*_450_ ≥0·175)) and 9 out of 311 participants seroreverted (i.e. loss of reactivity against either spike (*A*_450_ <0·175) or NCP (*A*_450_ <0·1905)).

### Adjusted Seroprevalence estimates

Bayesian multilevel regression estimated the prevalence of SARS-CoV-2 antibodies in our local HCW population at the initial blood draw (see Tables S3 to S7 for model parameters and response variables used in each model). The overall adjusted seroprevalence in the cohort was 23·1% (95% CrI 14·1–33·3), but varied across job type, job location and COVID-19 zone (Fig. 1). For job type, there was relatively high seroprevalence in occupational and physiotherapists of 39·2% (95% CrI 24·4–56·5), and relatively low seroprevalence in allied medical staff of 9·2% (95% CrI 1·4–21·3). Between wards, there was substantially higher seroprevalence in the AMU of 41·1% (95% CrI 30·0–52·9) compared to other wards. Across COVID-19 zones, working in the areas with the highest degree of COVID-19 patient contact (zone 6) saw a slightly higher seroprevalence of 28·6% (95% CrI 24·0–33·5) compared to the other five groups (Fig. 1). The adjusted proportion of asymptomatic cases was 38·9% (95% CrI 23·6–57·3) (Fig. S5). The proportion of asymptomatic cases remains relatively consistent across all covariates except for job type, where it ranges from as low as 21·4% for occupational and physiotherapists up to 61·5% for those who work in domestic services (Fig. S5).

**Figure 1.**
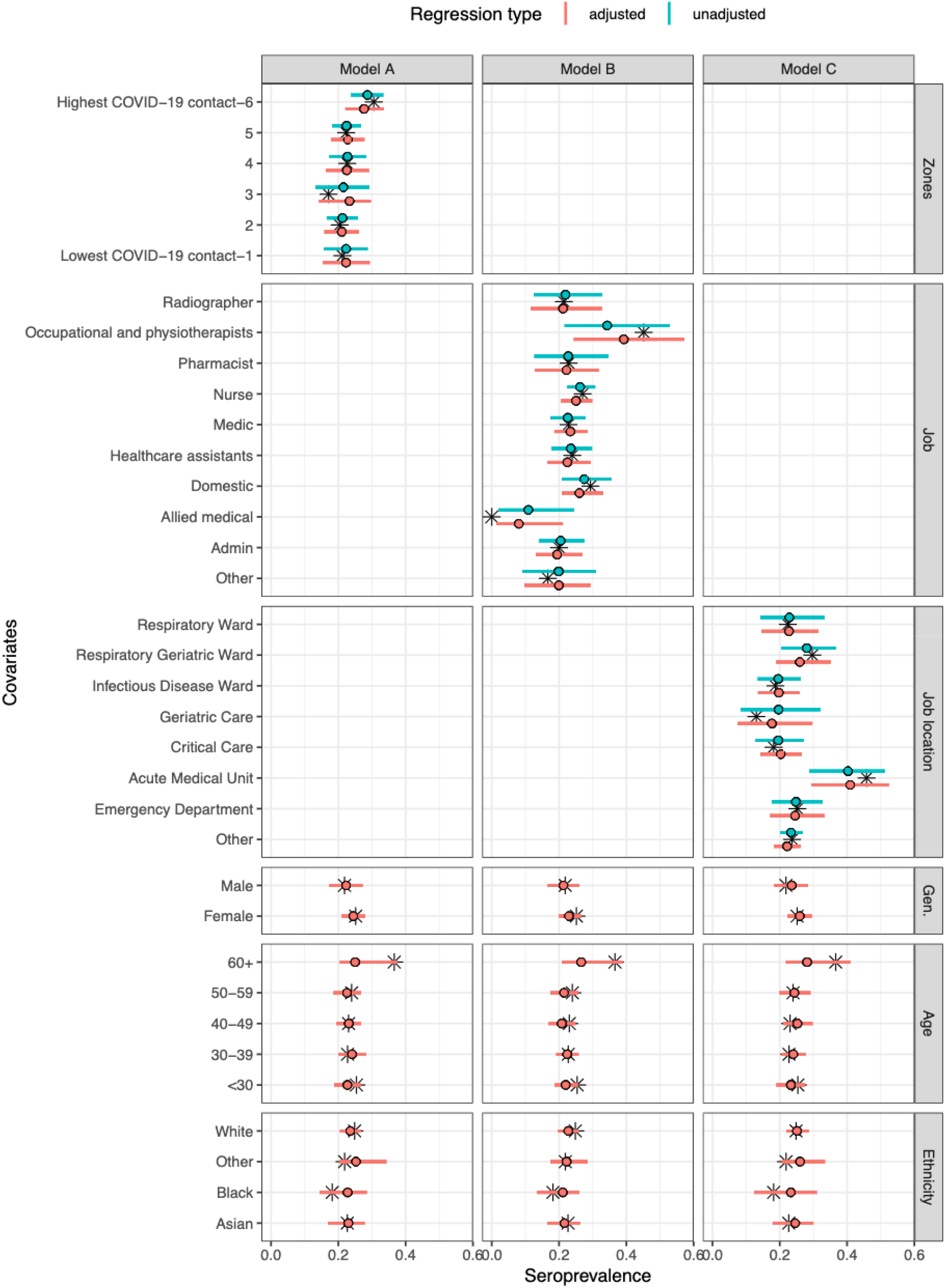
Model-predicted seroprevalence estimates for three different models (A-C), adjusted and unadjusted with covariates gender, age group, and ethnicity. Black stars represent point values from the data. The point and whiskers represent the mean value and 95% CrI of the posterior distribution. The three models differed by their primary exposure, Model A used COVID-19 zones (defined in supplementary information, 1 refers to lowest COVID-19 contact and 6 refers to highest COVID-19 contact), Model B the job role, and Model C the job location. Each model was evaluated either unadjusted (primary exposure only) or adjusted (primary exposure with age, gender, and ethnicity).

### Antibody kinetics model

Antibody concentration in log2 antibody units (logAU) were estimated for each study time point and the difference in antibody concentration between samples, for four different antibody-antigen interactions (spike-IgG, NCP-IgG, spike-IgA, NCP-IgA) were calculated. Though there was a positive correlation between Spike-IgG and NCP-IgG across all samples (R^2^= 0·53), the correlations between serum IgG and IgA were much weaker (R^2^ between 0·17 and 0·3) (Fig. S6).

The median estimated antibody titre at the first blood draw for spike-IgG was 6·7 (95% CrI 6·5–6·9) and for NCP-IgG was 6·2 (95% CrI 6·0–6·4) across all samples but varied by age group, gender, ethnicity and disease severity (symptomatic or asymptomatic) (Fig. 2). For both serum IgG and IgA, older age groups showed higher antibody titres. For example, the median log2 titre of spike-IgG at ≤30 years of age was 6·6 (95% CrI 6·2–7·0), and at 60+ years was 7·1 (95% CrI 6·6–7·8), while spike-IgA titre at ≤30 years of age was 6·8 (95% CrI 6·3– 7·3), and at 60+ years was 7·6 (95% CrI 7·0–8·3). Symptomatic cases showed similar titres compared to asymptomatic cases across IgG and IgA-serum measures (e.g. spike-IgG asymptomatic titre was 7·1 (95% CrI 6·7–7·4), and symptomatic titre was 7·2 (95% CrI 7·0–7·5)). Though there was a notable variation in estimated antibody titre across different ethnic groups, the small size of the Black and Asian participant sample resulted in large, overlapping credible intervals, making interpretation difficult. The reduction in antibody titre at the second blood draw was less in the spike-IgG (mean −0·15 (95% CrI −0·25– −0·06)) compared to NCP-IgG (mean −0·49 (95% CrI −0·58– −0·540)) and Spike-IgA/NCP-IgA. These estimated rates of decline remained consistent across all covariate groups studied. Consequently, the estimated time until seroreversion for seropositive samples from symptomatic participants was around 100 weeks for the spike-IgG, and 52 weeks for NCP-IgG and IgA serum measures (Fig. 2).). When considering seropositive samples from symptomatic participants across the four different antibody-antigens measures, there was little difference in the decrease in antibody levels as time post-symptom onset increased (Fig. S7).

**Figure 2.**
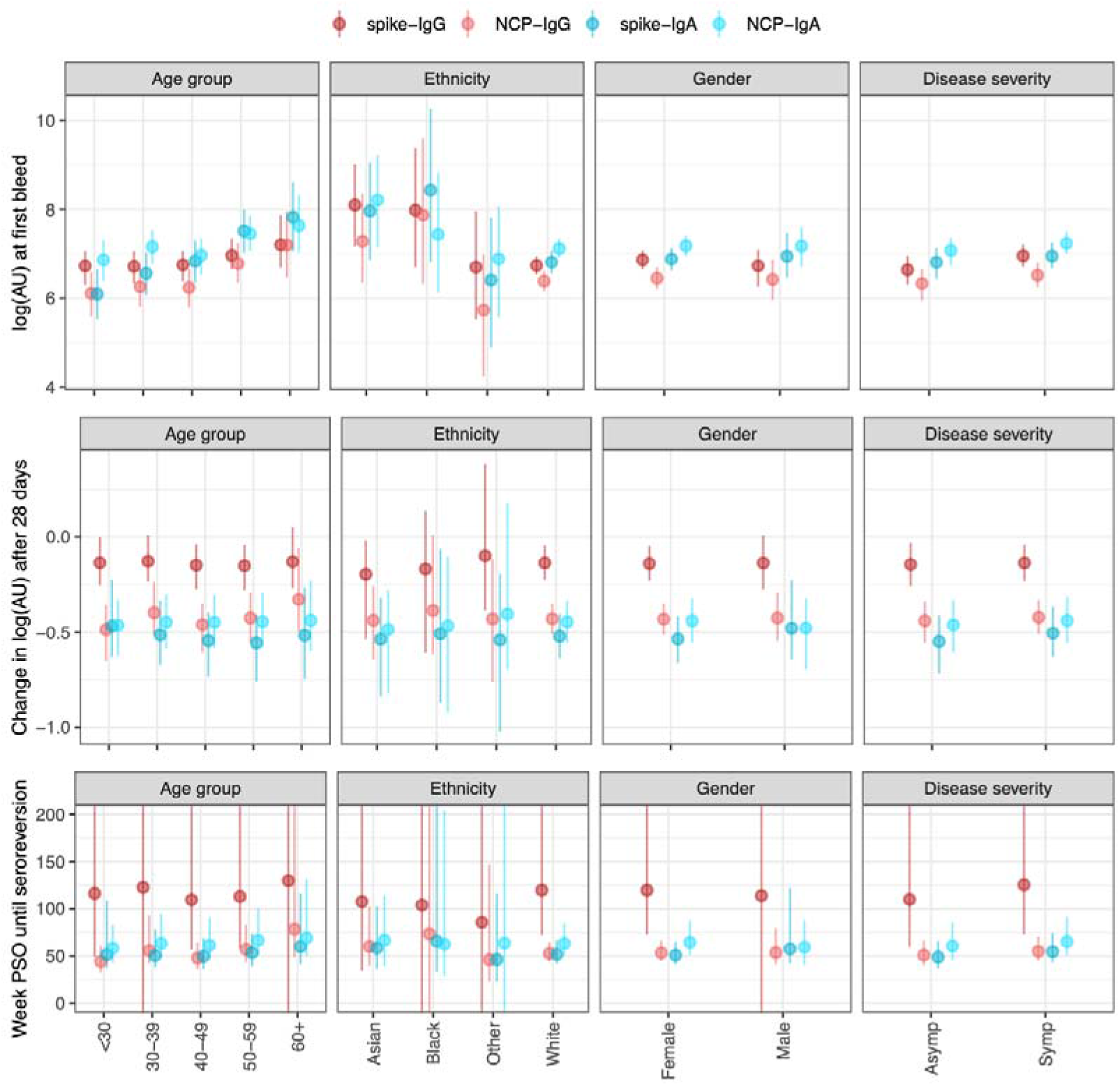
Outputs from the antibody kinetics model for four antibody-antigen interactions (spike-IgG, NCP-IgG, spike-IgA, and NCP-IgA). The IgG measures are in the WHO standard universal log2 antibody units, whereas the IgA measures are in log2(AU) units scaled relative to the values in the study. The dots show the median and the line segments show the 95% credible interval of the posterior distributions. Top panel shows the log2(AU) at the first bleed across four different covariates (Age group, ethnicity, gender, and disease severity). Middle panels show the change in log2(AU) after 30 days. The bottom panels show the time until seroreversion in weeks. Asymp (asymptomatic participants), Symp (symptomatic participants) PSO (post symptom onset).

### Heterogeneous sensitivity model

The heterogeneous sensitivity model demonstrates that using varying *A*_450_ cut-offs (corresponding to specific chosen sensitivity values from the assay validation dataset) to categorise seropositivity in the HERO dataset will result in a lower sensitivity than that we defined using our assay validation dataset (Fig. 3a). This is a consequence of the difference in the distributions of *A*_450_ values between the individuals included in the assay validation dataset (consisting of convalescent sera from SARS-CoV-2 NAAT-confirmed individuals), and the individuals included in the HERO dataset. These distribution differences mean that when the *A*_450_ cut-off value for a given sensitivity in the assay validation dataset is used to classify seropositivity in the HERO dataset, there is a greater proportion of false negatives. The model also shows that there is no difference in implied sensitivity between using spike or NCP as the antigenic target in the ELISA assay.

**Figure 3.**
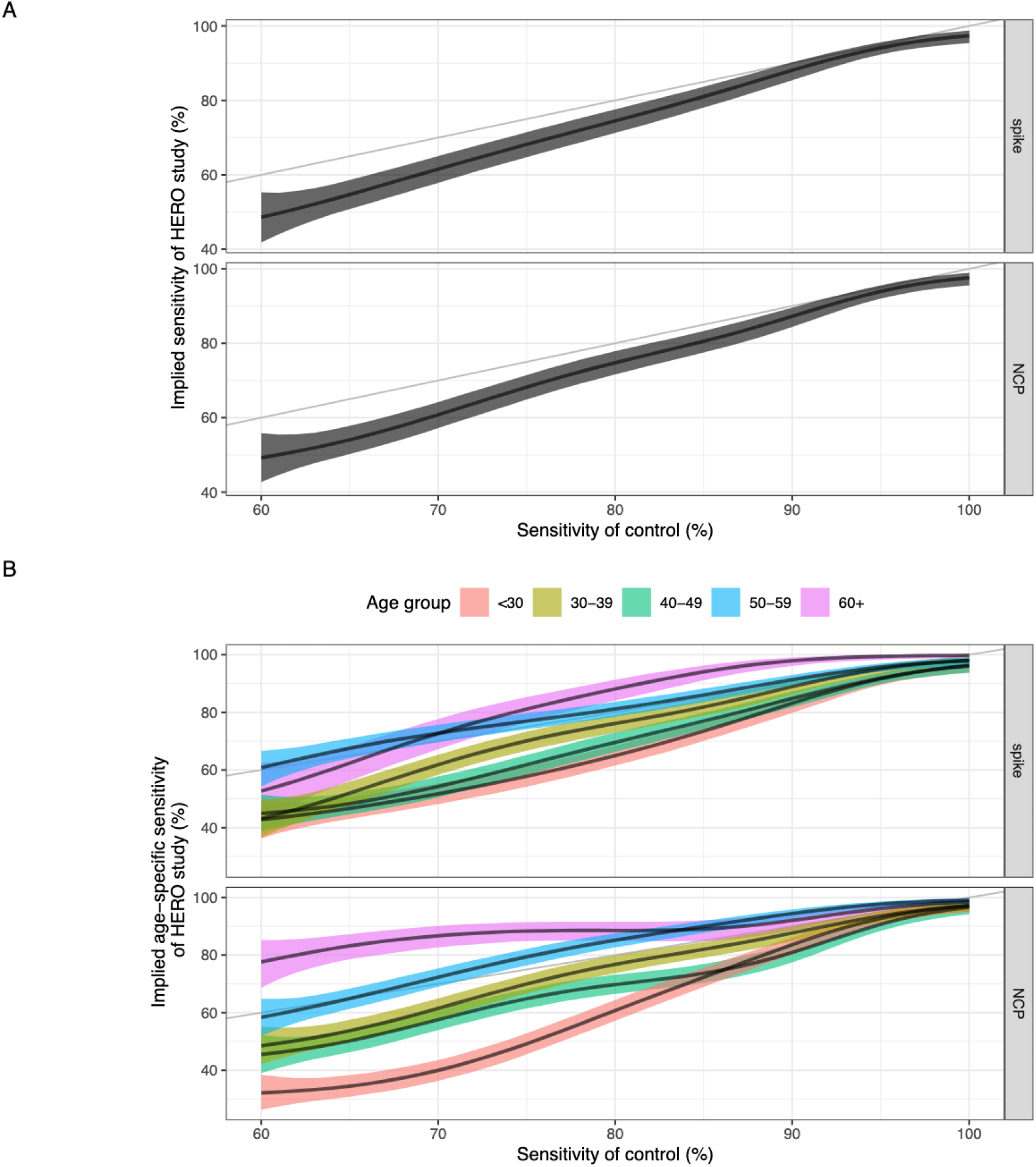
(a) Sensitivity of the assay validation dataset against the implied sensitivity of the HERO dataset for spike and nucleoprotein. (b) Sensitivity of the assay validation dataset against the implied age-specific sensitivity in the HERO dataset for spike and nucleoprotein. Black line and ribbon shows median and 95% CrI for the posterior distributions respectively.

The relationship between the *A*_450_ cut-off value and the sensitivity and specificity for the assay validation datasets for each antigen were plotted with the associated ROC curves (Figs. S8, S9). Given the higher antibody titres seen in older adults, we hypothesised that the corresponding higher *A*_450_ values suggest that commercial serological assays may have a higher sensitivity in detecting COVID-19 antibodies in older age groups compared with younger age groups. We therefore used the heterogenous sensitivity model to estimate age-specific implied sensitivity for assays of different sensitivity profiles to detect antibodies in the HERO dataset. We found that the sensitivity of a serological assay decreases with age due to the higher antibody titres seen in older people, with a clearer trend in an NCP-based assay compared to a spike-based assay (Fig 3b.). Assuming a theoretical assay validation set sample sensitivity of 80% for the NCP protein, the resulting median implied sensitivity for age groups <30, 30–39, 40–49, 50–59, and 60+ years was 61%, 77%, 70%, 85%, and 89% respectively.

## DISCUSSION

We found a high seroprevalence of SARS-CoV-2 in HCWs at a large UK hospital trust compared to national seroprevalence estimates, following the first pandemic wave in the UK (from March 2020 to June 2020).^26^ In addition, we identified important risk factors associated with occupational exposure to COVID-19, and described a significant correlation between age and the likelihood of a positive serological result which has important implications for the validation of SARS-CoV-2 antibody assays and the hitherto interpretation of population-level COVID-19 serology data.

Over 20% of HCWs at STH had evidence of SARS-CoV-2 infection within just over 100 days of the first confirmed COVID-19 patient being admitted to our NHS trust. This high proportion over a short space of time is likely representative of the much higher exposure to SARS-CoV-2 infection among certain subpopulations of the workforce that we tested, specifically HCWs in a patient facing role. Although data from other settings and countries suggested infection risk in HCWs is similar to community exposure, this seroprevalence is much higher than estimated seropositivity in the UK population at a similar time (6·0% (95 CrI 5·8–6·1) for July 2020).^26,27^

Our data show that HCWs working in AMUs are at significantly higher risk of infection with SARS-CoV-2, with seropositivity rates above that of other wards, and consistent with other UK studies.^5,6,28^ EDs face a similar patient turnover yet have lower HCW seroprevalence rates in both ours and previous reports.^5,6,28,29^ Although there are some patient factors which may increase HCW risk of infection from patients on AMU compared to ED (cohort bays, longer stays, more fomites e.g. bedside tables, chairs), transmission between HCWs may play a significant role. Universal surgical mask wearing in all hospital areas at STH was not mandated until after the first blood draw had occurred in our study, which would have allowed transmission between staff, particularly as a large proportion of seropositive HCWs were asymptomatic (38·3%) and therefore not self-isolating. HCWs working on AMU will also be more likely to interact with larger numbers of other HCWs at nurses’ stations, in doctors’ offices and break rooms (e.g. rotating on call medics, speciality medic and nursing teams, rapid response physiotherapy and occupational therapy), again likely further facilitating transmission amongst HCWs. In the event of a further wave or outbreak, infection, prevention and control (IPC) interventions to reduce the risk of individual occupational exposure in these areas could include targeted IPC training and auditing (particularly of PPE use and break areas), serial staff testing, pop-up isolation units in bay areas and optimising staff to patient ratios. Reviewing rotas to keep staff in smaller bubbles would be ideal but difficult to accommodate with current NHS workforce numbers. At the opposite end of the spectrum, HCWs in critical care units have some of the lowest seropositivity rates, which likely reflects ‘Level 3 Airborne’ PPE use on our Critical Care unit from early on in the pandemic (as opposed to ‘Level 2 Droplet’ PPE in other inpatient areas) plus increased availability of negative pressure rooms due to the widespread use of AGPs. This signal has been seen consistently in other seroprevalence studies.^5,6,8,28,29^

Occupational and physiotherapists (OT/PT) had the highest rates of seroprevalence across job roles in our cohort, with 45·5% seropositive on testing and modelling, which is consistent with some other UK studies.^6,30^ OT/PT both have prolonged close contact with patients whilst doing assessments, and in addition PT perform chest physiotherapy and open suctioning of the respiratory tract. Performing AGPs wearing ‘Level 2 Droplet’ PPE could result in acquisition of COVID-19 through aerosol inhalation. We could also speculate that as OT/PT work across several different inpatient areas, and then mix in outpatient therapy departments, this could increase risk of infection from both patients and other HCWs.

Increasing age was associated with seropositivity, with over a third of our HCWs aged over 60 testing seropositive, and with higher antibody titres. As those who were older had higher SARS-CoV-2 IgG and IgA titres, we hypothesised that this may lead to variable age-specific assay performance where an assay’s sensitivity was low. Our findings show that the sensitivity of a serological assay increases with increasing age due to the higher antibody titres seen in older people, with a clearer trend in an NCP-based assay compared to a spike-based assay. Consistent with other studies, we also found that NCP-IgG is likely to wane more quickly than Spike-IgG. Serological testing based on NCP-IgG alone may therefore underestimate seroprevalence, especially as time increases from major pandemic waves. As two of the major commercial SARS-CoV-2 antibody assays (Roche Elecsys and Abbott SARS-CoV-2 IgG) validated their assay using serum from patients with more severe disease early on in the pandemic (i.e. those who presented to health services), it would be reasonable to assume these cases were also primarily older in age.^31,32^ Previous studies have also shown that sera from patients with severe COVID-19 have higher antibody titres.^33,34^ Our heterogeneous sensitivity model suggests that using samples from severe symptomatic cases in older people to calibrate serological assays results in lower sensitivity when used to classify milder, community cases which would further underestimate seroprevalence. In addition, calibrating ELISAs on datasets containing a large proportion of samples collected from older age groups of patients provides lower sensitivity when applied to datasets with a smaller proportion of samples collected from older age groups, where the log2AU units are much closer to the *A*_450_ cut-off (Fig. S10). Using an assay which detects spike IgG, that has been validated using serum samples from a broader population, with antibody kinetics modelling and/or with age adjusted cut offs could overcome these limitations. However, with increasing vaccine coverage, use of spike IgG to determine seroprevalence becomes more problematic when distinguishing whether an individual is seropositive from vaccination or previous infection. Assays which combine antibody responses to membrane protein with NCP antibodies may overcome these challenges.^35,36^

We note the limitations of our study, which include a potential for selection bias due to participants self-enrolling for convenience, rather than using systematic sampling. Reassuringly, our seroprevalence rates are similar to those seen in other UK based seroprevalence studies.^5,30^ In addition, we recognise that our cohort has relatively low numbers of HCWs from minority ethnic backgrounds (∼10%), compared to the Sheffield general population (19%).^37^

With the ongoing global devastation due to the COVID-19 pandemic, knowledge of HCW exposure risk factors is vital, given the possibility of reinfection with immune-escape variants, and that future re-vaccination programmes are likely on the horizon. Our real-world data demonstrate that HCWs are at high risk of exposure, particularly those who work on AMU or as PT/OT as well as domestic services personnel. Measuring and correctly interpreting population seroprevalence data can help guide decision makers on risk management, so access to a serological assay that is both sensitive and specific is critical. Our data also emphasise the relationship between increasing age and greater antibody titres is key to accurate interpretation of serostatus.

## Data Availability

An R package containing all the analysis in this study is available at https://github.com/dchodge/hero-study.

## Acknowledgements

We would like to thank all the participants who generously donated their time for this study. We would like to thank the technician team from the Department of Infection, Immunity and Cardiovascular Disease (Mark Ariaans, Yvonne Stephenson, Linda Kay, Ben Durham, Amy Lewis, Barbora Ndreca) and Department of Oncology and Metabolism (Julie Porter, Maggie Glover and Ana Lopez) for help with processing samples, as well as Amanda Blythe, Julie Schofield, Phil Ravencroft, Elizabeth Kirkssen, Katie Jenkins, Sarah Berry, Debbie Allen and Aimee Card for help with participant recruitment, sampling and regulatory support. We thank Professor William Egner and Dr Ravi Sargur for useful discussions on serological assay validation. We are grateful to Professor Florian Krammer, Fatima Amanat and Parnavi Desai for the provision of the spike constructs.

## References

1 Nguyen Long H, Drew David A, Graham Mark S, Joshi Amit D, Guo Chuan-Guo, Ma Wenjie, et al. Risk of {COVID-19} among front-line health-care workers and the general community: a prospective cohort study. Lancet Public Heal 2020;5(9):e475--e483.

2 Mutambudzi Miriam, Niedwiedz Claire, Macdonald Ewan Beaton, Leyland Alastair, Mair Frances, Anderson Jana, et al. Occupation and risk of severe COVID-19: prospective cohort study of 120 075 UK Biobank participants. Occup Environ Med 2020. Doi: 10.1136/oemed-2020-106731.

3 Behrens Georg M N, Cossmann Anne, Stankov Metodi V, Witte Torsten, Ernst Diana, Happle Christine, et al. Perceived versus proven SARS-CoV-2-specific immune responses in health-care professionals. Infection 2020;48(4):631–4. Doi: 10.1007/s15010-020-01461-0.

4 Keeley A.J., Evans C., Colton H., Ankcorn M., Cope A., State A., et al. Roll-out of SARS-CoV-2 testing for healthcare workers at a large NHS Foundation Trust in the United Kingdom, March 2020. Eurosurveillance 2020;25(14). Doi: 10.2807/1560-7917.ES.2020.25.14.2000433.

5 Shields Adrian, Faustini Sian E., Perez-Toledo Marisol, Jossi Sian, Aldera Erin, Allen Joel D., et al. SARS-CoV-2 seroprevalence and asymptomatic viral carriage in healthcare workers: A cross-sectional study. Thorax 2020. Doi: 10.1136/thoraxjnl-2020-215414.

6 Eyre David W., Lumley Sheila F., O’donnell Denise, Campbell Mark, Sims Elizabeth, Lawson Elaine, et al. Differential occupational risks to healthcare workers from SARS-CoV-2 observed during a prospective observational study. Elife 2020. Doi: 10.7554/ELIFE.60675.

7 Cooper Daniel J., Lear Sara, Watson Laura, Shaw Ashley, Ferris Mark, Doffinger Rainer, et al. A prospective study of risk factors associated with seroprevalence of SARS-CoV-2 antibodies in healthcare workers at a large UK teaching hospital. MedRxiv 2020. Doi: 10.1101/2020.11.03.20220699.

8 Valdes Ana M, Moon James C, Vijay Amrita, Chaturvedi Nish, Norrish Alan, Ikram Adeel, et al. Longitudinal assessment of symptoms and risk of SARS-CoV-2 infection in healthcare workers across 5 hospitals to understand ethnic differences in infection risk. EClinicalMedicine 2021:100835. Doi: 10.1016/j.eclinm.2021.100835.

9 Hou Hongyan, Wang Ting, Zhang Bo, Luo Ying, Mao Lie, Wang Feng, et al. Detection of IgM and IgG antibodies in patients with coronavirus disease 2019. Clin Transl Immunol 2020. Doi: 10.1002/cti2.1136.

10 Deeks Jonathan J., Dinnes Jacqueline, Takwoingi Yemisi, Davenport Clare, Spijker René, Taylor-Phillips Sian, et al. Antibody tests for identification of current and past infection with SARS-CoV-2. Cochrane Database Syst Rev 2020. Doi: 10.1002/14651858.CD013652.

11 Burbelo Peter D., Riedo Francis X., Morishima Chihiro, Rawlings Stephen, Smith Davey, Das Sanchita, et al. Detection of nucleocapsid antibody to SARS-CoV-2 is more sensitive than antibody to spike protein in COVID-19 patients. MedRxiv 2020. Doi: 10.1101/2020.04.20.20071423.

12 Ainsworth Mark, Andersson Monique, Auckland Kathryn, Baillie J. Kenneth, Barnes Eleanor, Beer Sally, et al. Performance characteristics of five immunoassays for SARS-CoV-2: a head-to-head benchmark comparison. Lancet Infect Dis 2020. Doi: 10.1016/S1473-3099(20)30634-4.

13 Reynolds Catherine J., Swadling Leo, Gibbons Joseph M., Pade Corinna, Jensen Melanie P., Diniz Mariana O., et al. Discordant neutralizing antibody and T cell responses in asymptomatic and mild SARS-CoV-2 infection. Sci Immunol 2020. Doi: 10.1126/SCIIMMUNOL.ABF3698.

14 Grandjean Louis, Saso Anja, Torres Arturo, Lam Tanya, Hatcher James, Thistlethwayte Rosie, et al. Humoral response dynamics following infection with SARS-CoV-2. MedRxiv 2020. Doi: 10.1101/2020.07.16.20155663.

15 Wang Yanan, Li Jingjing, Li Huijun, Lei Ping, Shen Guanxin, Yang Chunguang. Persistence of SARS-CoV-2-specific antibodies in COVID-19 patients. Int Immunopharmacol 2021. Doi: 10.1016/j.intimp.2020.107271.

16 Wu Fan, Wang Aojie, Liu Mei, Wang Qimin, Chen Jun, Xia Shuai, et al. Neutralizing antibody responses to SARS-CoV-2 in a COVID-19 recovered patient cohort and their implications. MedRxiv 2020. Doi: 10.1101/2020.03.30.20047365.

17 Orner Erika P., Rodgers Mary A., Hock Karl, Tang Mei San, Taylor Russell, Gardiner Mary, et al. Comparison of SARS-CoV-2 IgM and IgG seroconversion profiles among hospitalized patients in two US cities. Diagn Microbiol Infect Dis 2021. Doi: 10.1016/j.diagmicrobio.2020.115300.

18 Gorse Geoffrey J., Donovan Mary M., Patel Gira B. Antibodies to coronaviruses are higher in older compared with younger adults and binding antibodies are more sensitive than neutralizing antibodies in identifying coronavirus-associated illnesses. J Med Virol 2020. Doi: 10.1002/jmv.25715.

19 Ho Mei Shang, Chen Wei Ju, Chen Hour Young, Lin Szu Fong, Wang Win Chin, Di Jiali, et al. Neutralizing antibody response and SARS severity. Emerg Infect Dis 2005. Doi: 10.3201/eid1111.040659.

20 CQC. Sheffield Teaching Hospitals CQC Report 2018. 2018.

21 Amanat Fatima, Stadlbauer Daniel, Strohmeier Shirin, Nguyen Thi H.O., Chromikova Veronika, McMahon Meagan, et al. A serological assay to detect SARS-CoV-2 seroconversion in humans. Nat Med 2020. Doi: 10.1038/s41591-020-0913-5.

22 Johari Yusuf B., Jaffé Stephen R.P., Scarrott Joseph M., Johnson Abayomi O., Mozzanino Théo, Pohle Thilo H., et al. Production of trimeric SARS-CoV-2 spike protein by CHO cells for serological COVID-19 testing. Biotechnol Bioeng 2021. Doi: 10.1002/bit.27615.

23 Tugaeva Kristina V, Hawkins Dorothy E D P, Smith Jake L R, Bayfield Oliver W, Ker De-Sheng, Sysoev Andrey A, et al. The Mechanism of SARS-CoV-2 Nucleocapsid Protein Recognition by the Human 14-3-3 Proteins. J Mol Biol 2021:166875.

24 Flaxman Seth, Mishra Swapnil, Gandy Axel, Unwin H. Juliette T., Mellan Thomas A., Coupland Helen, et al. Estimating the effects of non-pharmaceutical interventions on COVID-19 in Europe. Nature 2020. Doi: 10.1038/s41586-020-2405-7.

25 Hanson Kimberly E, Caliendo Angela M, Arias Cesar A, Englund Janet A, Hayden Mary K, Lee Mark J, et al. Infectious Diseases Society of America Guidelines on the Diagnosis of COVID-19: Serologic Testing. [WwwIdsocietyOrg/COVID19guidelines/Serology](http://www.IdsocietyOrg/COVID19guidelines/Serology) 2020.

26 Ward Helen, Cooke Graham, Atchison Christina, Whitaker Matthew, Elliott Joshua, Moshe Maya, et al. Declining prevalence of antibody positivity to SARS-CoV-2: A community study of 365,000 adults. MedRxiv 2020. Doi: 10.1101/2020.10.26.20219725.

27 Steensels Deborah, Oris Els, Coninx Laura, Nuyens Dieter, Delforge Marie Luce, Vermeersch Pieter, et al. Hospital-Wide SARS-CoV-2 Antibody Screening in 3056 Staff in a Tertiary Center in Belgium. JAMA - J Am Med Assoc 2020. Doi: 10.1001/jama.2020.11160.

28 Houlihan Catherine F., Vora Nina, Byrne Thomas, Lewer Dan, Kelly Gavin, Heaney Judith, et al. Pandemic peak SARS-CoV-2 infection and seroconversion rates in London frontline health-care workers. Lancet 2020. Doi: 10.1016/S0140-6736(20)31484-7.

29 Galán M. A. Isabel, Velasco María, Casas M. A. Luisa, Goyanes M. A. José, Rodríguez-Caravaca Gil, Losa Juan Emilio, et al. SARS-CoV-2 Seroprevalence among all workers in a teaching hospital in Spain: Unmasking the risk. MedRxiv 2020. Doi: 10.1101/2020.05.29.20116731.

30 El Bouzidi Kate, Pirani Tasneem, Rosadas Carolina, Ijaz Samreen, Pearn Matthew, Chaudhry Shehnila, et al. Severe Acute Respiratory Syndrome Coronavirus-2 Infections in Critical Care Staff: Beware the Risks beyond the Bedside. Crit Care Med 2021. Doi: 10.1097/CCM.0000000000004878.

31 Public Health England. Evaluation of Roche Elecsys Anti-SARS-CoV-2 serology assay for the detection of anti-SARS-CoV-2 antibodies About Public Health England 2020.

32 Public Health England. Evaluation of the Abbott SARS-CoV-2 IgG for the detection of anti-SARS-CoV-2 antibodies About Public Health England 2020:1–19.

33 Garcia-Beltran Wilfredo F., Lam Evan C., Astudillo Michael G., Yang Diane, Miller Tyler E., Feldman Jared, et al. COVID-19-neutralizing antibodies predict disease severity and survival. Cell 2021. Doi: 10.1016/j.cell.2020.12.015.

34 Lau Eric H.Y., Tsang Owen T.Y., Hui David S.C., Kwan Mike Y.W., Chan Wai hung, Chiu Susan S., et al. Neutralizing antibody titres in SARS-CoV-2 infections. Nat Commun 2021. Doi: 10.1038/s41467-020-20247-4.

35 Bruckner Tim A., Parker Daniel M., Bartell Scott M., Vieira Veronica M., Khan Saahir, Noymer Andrew, et al. Estimated seroprevalence of SARS-CoV-2 antibodies among adults in Orange County, California. Sci Rep 2021. Doi: 10.1038/s41598-021-82662-x.

36 Martin Sophie, Heslan Christopher, Jégou Gwénaële, Eriksson Leif A., Le Gallo Matthieu, Thibault Vincent, et al. SARS-CoV2 Envelop Proteins Reshape the Serological Responses of COVID-19 Patients. SSRN Electron J 2021. Doi: 10.2139/ssrn.3821960.

37 Sheffield City Council. Briefing Note 2□: Ethnicity. 2012.

