## Supplementary Materials for "Risk factors for SARS-CoV-2 seroprevalence following the first pandemic wave in UK healthcare workers in a large NHS Foundation Trust"

**Supplementary Information**

### Levels of COVID-19 PPE used at STH

‘Level 3 Airborne’ used for High Consequence Infectious Diseases: Triple gloved, long sleeved fluid resistant gown, thick plastic apron, hood, visor, FFP3 mask, wellies

‘Level 2 Airborne’ used in aerosol generating procedure (AGP) COVID-19 areas: Single gloved, long sleeved fluid resistant gown, FFP3 mask, visor

‘Level 2 Droplet’, as used for routine care of COVID-19: Single gloved, plastic apron, surgical mask (+/- visor if splash risk)

### COVID-19 Zones

Red Zones are defined as areas caring for patients with suspected / confirmed COVID-19

Green Zones are defined as areas caring for patients who do not have suspected / confirmed COVID-19

Risk stratification:

1 = I've not worked in any patient areas (lowest COVID-19 contact)

2 = I've worked in green zones most days

3 = I've attended/cared for COVID-19 patients but only after they've been declared no longer infectious

4 = I work in red zones most days but don't attend/care for patients myself

5 = I've attended/cared for patients with COVID-19 in red zones occasionally

6 = I've attended/cared for patients with COVID-19 in red zones most days (highest COVID-19 contact)

### Stratification of COVID-19 diagnosis during the first wave of the UK pandemic

Study participants were asked to recall if they had been unwell with a COVID-19 consistent illness since the beginning of February 2020. The certainty of an illness being consistent with COVID-19 was classified as:

- Diagnosed COVID-19 and NAAT confirmed
  (“YES - I was diagnosed and it was confirmed with a swab test”),
- Diagnosed COVID-19 but NAAT not performed
  (“YES - I was diagnosed but it has not been confirmed with swab test”),
- Self reported Symptomatic
  (“MAYBE - I've had symptoms of cough, fever or headache or required days of work for a new medical problem”),
- Asymptomatic
  (“NO” or “NO and I’ve had a negative PCR test”)

### Indirect ELISA to detect anti-SARS-CoV-2 haemagglutinin IgG in human serum samples - method validation

#### Coating concentrations

Plate coating concentrations of SARS-CoV-2 spike and nucleoprotein (NCP) were determined via checkerboarding with SARS-CoV-2 positive convalescent sera from individuals with nucleic acid amplification test (NAAT) confirmed SARS-CoV-2 and pre-pandemic negative sera. Concentrations were chosen to maximise sensitivity while minimising non-specific binding. For the initial batches of protein used to set up the assay, 2 µg/mL was selected as the optimal coating concentration for both spike and NCP. Subsequent batches of protein were validated via checkerboarding against control sera, and a concentration giving Absorbance450 (*A*_450_) closest to those obtained with the original batches (within 10% coefficient of variance (CV)) was chosen.

#### Secondary antibody concentrations

The assay dilution of the secondary antibody (goat anti-human IgG-HRP conjugate) was again determined via checkerboarding with NAAT+ and pre-pandemic sera, and selected to maximise signal while avoiding an increase in non-specific binding. Increasing the concentration of the secondary antibody resulted in a significant increase in signal, with little to no increase in background, and so 1:500 was selected as the optimal dilution for the assay.

#### Sample dilutions

Sample screening dilutions were selected by serially diluting sera from both the NAAT+ and pre-pandemic sets from 1:50 to 1:250, and comparing the *A*_450_ of the two. 1:200 was selected as the optimal dilution for both spike and NCP as it reduced much of the non-specific binding seen at higher concentrations in the pre-pandemic samples, while still allowing NAAT+ samples to achieve good signal.

#### Setting Thresholds for seropositivity

We ran a validation panel of 190 samples from SARS-CoV-2 NAAT-confirmed cases (at least 14 days from positive test) and 675 pre-pandemic samples (taken prior to 2017) on both proteins, and used receiver operating characteristic (ROC) curves to set thresholds. Positive validation samples came from both inpatients with severe disease and outpatients with mild or no symptoms. Guidelines published in August 2020 by the Infectious Diseases Society of America recommend that serologic assays to detect anti-SARS-CoV-2 antibodies should have high specificity (≥99.5%), especially where seroprevalence in the community is expected to be low.^1^ Although individually both the spike and NCP assays are highly sensitive, neither achieve this 99.5% specificity alone. In order to achieve both high sensitivity and specificity when assigning serostatus, we chose individual assay thresholds which maximised sensitivity, and defined a SARS-CoV-2 seropositive sample as one where both the spike and NCP were above threshold to achieve high specificity. This allows us to achieve sensitivity of 99.47% and specificity of 99.56%.

##### Spike assay alone

| **Spike threshold** | **Sensitivity** | **95% CI** | **Specificity** | **95% CI** |
| --- | --- | --- | --- | --- |
| 0.1753 | 99.47% | 97.08% to 99.97% | 98.81% | 98.68% to 99.40% |

##### NCP assay alone

| **NCP threshold** | **Sensitivity** | **95% CI** | **Specificity** | **95% CI** |
| --- | --- | --- | --- | --- |
| 0.1905 | 99.47% | 97.08% to 99.97% | 84.15% | 81.20% to 86.71% |

##### Definition of seropositivity requiring both spike and NCP assays to be above threshold

| **Spike threshold** | **NCP threshold** | **Sensitivity** | **95% CI** | **Specificity** | **95% CI** |
| --- | --- | --- | --- | --- | --- |
| 0.1753 | 0.1905 | 99.47% | 97.10% to 99.99% | 99.56% | 98.71% to 99.91% |

### Table S1. Details of the samples used to set thresholds during assay validation.

Seropositivity was defined as a sample showing reactivity above thresholds set in both spike and NCP assays. Seropositive samples were taken at least 14 days from SARS-CoV-2 NAAT-positive test.

|  | **n** | **Seropositive** | **Seronegative** |
| --- | --- | --- | --- |
| **SARS-CoV-2 NAAT positive** | **190** | **189** | **1** |
| Inpatient | 52 | 52 | 0 |
| Outpatient | 138 | 137 | 1 |
| **Pre-pandemic sera collected prior to 2017** | **675** | **3** | **672** |
| Seasonal coronavirus positive (by NAAT) | 20 | 0 | 20 |
| Human Immunodeficiency Virus 1 positive | 14 | 0 | 14 |
| Dialysis patient | 9 | 0 | 9 |
| Cytomegalovirus IgM+ | 8 | 1 | 7 |
| Epstein-Barr Virus IgM+ | 5 | 0 | 5 |
| Influenza virus positive (by NAAT) | 6 | 0 | 6 |
| Rhinovirus positive (by NAAT) | 1 | 0 | 1 |
| Adenovirus positive (by NAAT) | 1 | 0 | 1 |
| Human metapneumovirus positive (by NAAT) | 1 | 0 | 1 |

### Figure S1. ROC curves of the spike and NCP assays

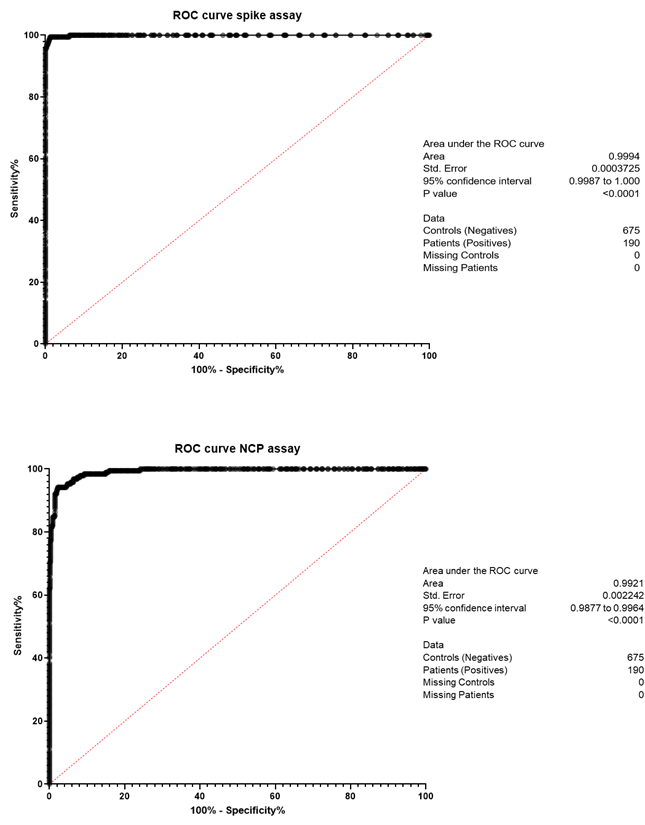

### Indirect ELISA to detect anti-SARS-CoV-2 haemagglutinin IgA in human serum samples

The IgA assay was validated in the same manner as the IgG assay. The spike and NCP protein coating concentrations were kept the same in both assays to maintain comparability of results. The optimum dilution of the secondary antibody (goat anti-human IgA-HRP conjugate) was determined to be 1:1000 for this assay, and 1:100 was selected as the optimum sample screening dilution.

We saw higher levels of cross-reactivity in the IgA assay than in the IgG assay, as seen in figure S2. Given this, and the fact that IgA levels decline more rapidly post-infection than IgG, it was difficult to set thresholds based on sensitivity and specificity for this assay. We opted instead to use the IgG assay alone to define seropositivity (as described above), and to use the IgA assay output as a way to compare the IgA response of seropositive participants only.

### Figure S2. Comparison of IgA assay *A*_450_ based on IgG Serostatus

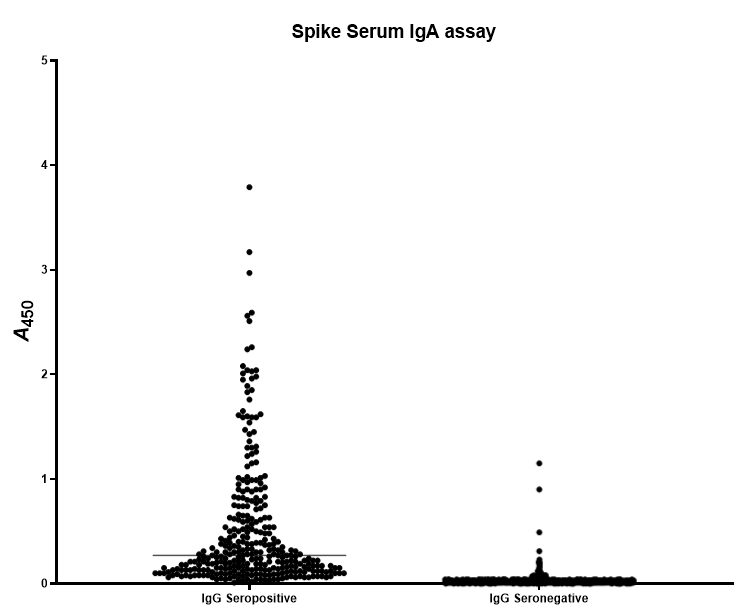

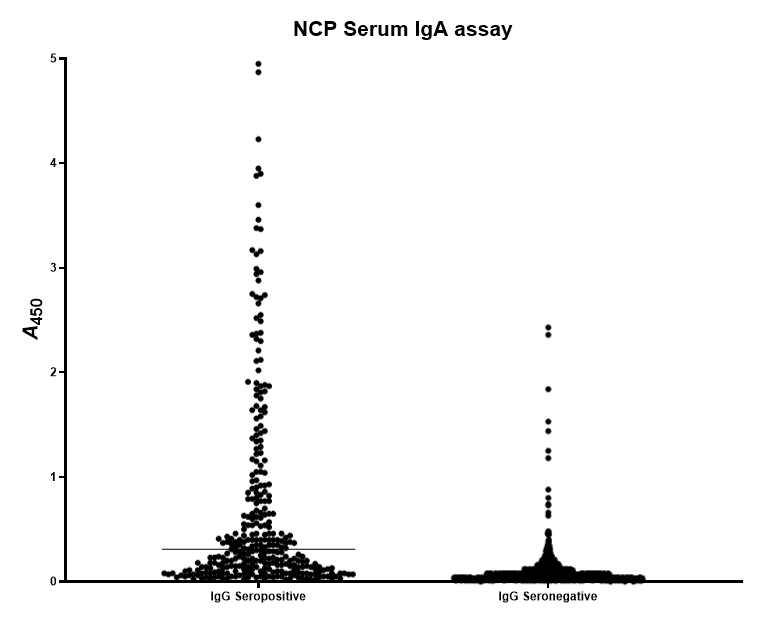

#### Calibration curve

In order to quantify the amount of antibody present in samples, a calibration curve was included on each plate and used to interpolate an antibody unit value for each sample *A*_450_. The calibration curve was created by pooling sera from two serum samples from NAAT+ve inpatients, chosen for their high *A*_450_ for both spike and NCP. The curve was generated by serially diluting from a starting concentration of 1:200 for IgG, or 1:100 for IgA (to match sample dilution), to create a 12-well dilution series. Following initial testing, 1.75× steps were chosen for the series in order to give a curve with the maximum number of points between saturation and threshold. An arbitrary value of 1000 antibody units was assigned to the most concentrated well of the series; the curve was then used to interpolate arbitrary antibody unit values for the samples on the plate.

#### Validation with WHO International Standard

For the IgG assay, following the manufacture of a WHO International Standard for anti-SARS-CoV-2 immunoglobulin (NIBSC, 20/136), our calibration curve was run in parallel with this Standard, as well as with the International reference panel (NIBSC 20/268).^2^ We assigned a WHO International Standard unit value to each dilution of our calibration curve (separately for spike and NCP). We are therefore able to interpolate WHO recommended IU/mL values for samples tested against our calibration curve. For a antigen protein *p*, and serial dilution *i*, we were able to convert arbitrary units, x^p^_i_, to WHO units, y^p^_i_, via the fitted linear equation:

y^S^_i_ = 0.0962 + x^S^_i_*1.0008, y^N^_i_ =0.6501 + x^N^_i_*0.934

with R^2^ = 0.0.9974 and R^2^ = 0.9984 for spike and NCP antigens respectively

### Table S2. Comparison of antibody units in assay calibration curve sera assigned to the assay with WHO international standard antibody units

| **Curve series** | **Serum dilution**  **(IgG)** | **Serum dilution**  **(IgA)** | **Arbitrary units** | **WHO units - Spike (IgG only)** | **WHO units - NCP (IgG only)** |
| --- | --- | --- | --- | --- | --- |
| 1 | 1:200 | 1:100 | 1000 | 922.74 | 976.32 |
| 2 | 1:350 | 1:175 | 571.43 | 527.28 | 557.90 |
| 3 | 1:612.50 | 1:306.25 | 326.53 | 301.30 | 318.80 |
| 4 | 1:1071.88 | 1:535.96 | 186.59 | 172.17 | 182.17 |
| 5 | 1:1875.78 | 1:937.89 | 106.62 | 98.38 | 104.10 |
| 6 | 1:3282.62 | 1:1641.31 | 60.93 | 56.22 | 59.48 |
| 7 | 1:5744.58 | 1:2872.29 | 34.82 | 32.13 | 33.99 |
| 8 | 1:10053.02 | 1:5026.51 | 19.89 | 18.36 | 19.42 |
| 9 | 1:17592.78 | 1:8796.39 | 11.37 | 10.49 | 11.10 |
| 10 | 1:30787.36 | 1:15393.68 | 6.50 | 5.99 | 6.34 |
| 11 | 1:53877.88 | 1:26938.94 | 3.71 | 3.43 | 3.62 |
| 12 | 1:94286.29 | 1:47143.14 | 2.12 | 1.96 | 2.07 |

#### Analysis

Results were analysed by custom program elisa-dolittle ([GitHub - lindseyb101112/elisa-dl](https://github.com/lindseyb101112/elisa-dl)). The amount of antibody present in participant samples was determined by interpolation against the calibration curve.

#### Quality control

Samples were tested in duplicate wells on ELISA plates. The maximum CV of duplicates was 10%; any sample duplicates with a CV >10% were retested. 2.8% of sample duplicates tested had >10% CV (n = 640). Median CV of sample duplicates was 2.0%.

Calibration curves were also tested in duplicate. Any calibration curve duplicates with a CV >10% were excluded from the generation of the curve.

Positive and negative controls were tested in triplicate. One well was excluded if CV across 3 wells was >10%, and this resulted in CV <10% for the remaining 2 wells. If no 2 wells were <10% CV, the plate was retested. The maximum CV of positive controls across plates was 20%; any plates with control values outside of this range were retested.

### Production of Nucleocapsid protein

A plasmid construct expressing untagged SARS-CoV-2 Nucleocapsid, (NCP, Nucleoprotein, Uniprot ID P0DTC9 (NCAP_SARS2) <https://www.uniprot.org/uniprot/P0DTC9>) under the control of a T7 promoter was supplied by Dr Fred Antson, University of York UK.^3^ The plasmid was transformed into in *Escherichia coli* BL21(DE3) cells carrying plasmid pRARE2 (obtained from Merck-Millipore, Catalog No. 71400) and plated on MDAG agar supplemented with chloramphenicol and carbenicillin at 34 and 100 mg/mL respectively at 37°C.^4^ A single colony was inoculated into 5 mL of liquid media as above and grown overnight. Cells were grown for 48 hrs in 500 mL of autoinducing media Super Broth Base (Formedium Ltd, UK) at 28°C from an initial inoculum of 1:1000 in a shaking incubator at 225 rpm in baffled 2.5 L flasks containing antibiotics as above. Samples were monitored for expression by SDS-PAGE electrophoresis. Cells were harvested by centrifugation, lysed and freed of contaminating nucleic acids essentially as described previously for T5 exonuclease *via* precipitation with polyethylenimine and the protein was concentrated by precipitation with ammonium sulphate.^5^ The protein pellet was resuspended in 50 mM HEPES/NaOH pH 8, 5% glycerol, 1 mM EDTA. Anionic contaminants were removed by passing the diluted protein through a prepacked 5 mL Q-Sepharose Fast Flow (Cytiva) and further purified by cation exchange chromatography on a 5 mL prepacked SP-Sepharose column (Cytiva). The column was washed with ~100 mL of the same buffer and 1 mL fractions were eluted with a 100 mL linear gradient containing 100 mM – 1 M NaCl. Protein fractions were pooled, concentrated with Amicon® Ultra-15 centrifugal filter units (Merck) to ~ 30 mg/mL. The protein was purified by size exclusion chromatography using an XK 16/100 column packed with Superdex 200 (Cytiva) eluted with 50 mM Tris-HCl pH 8.5, 5% glycerol, 1 mM EDTA and 1.0 M NaCl. Pooled fractions of pure protein were concentrated to ~50 mg/mL in Amicon® units and the buffer was exchanged to 50 mM Tris-HCl pH 8.5, 5% glycerol, 1 mM EDTA, 0.3 M NaCl. Purity was assessed by applying a dilution series on an SDS-PAGE gel and aliquots were stored at -80°C.

### Mathematical and Statistical Methods

#### Prevalence model A — Seroprevalence

##### Model outline

*Data*

We included all samples which included a serological sample at the first bleed (n = 1275).

*Model variables*

### Table S3. Summary of the response variables and the covariates used in the regression model.

The elements in the encoding vector are associated with the groups given in the categories column in the order they are written.

| **Variable** | **Description** | **Categories (order)** | **Symbol and encoding (respectively)** |
| --- | --- | --- | --- |
| *Response* | | | |
| Serostatus | Serostatus of individual | Seronegative, seropositive (2) | $y\in\{0, 1\}$ |
| *Covariates* | | | |
| Age | Age category of individual | <30, 30-39, 40-49, 50-59, 60+ years (5) | $x_{age}\in\{1, 2, 3, 4, 5\}$ |
| Ethnicity | Ethnicity of individual | White, Black, Asian, Other (4) | $x_{eth}\in\{1, 2, 3, 4\}$ |
| Gender | Gender of individual | Female , Male (2) | $x_{gen}\in\{1, 2\}$ |
| Job | Type of job individual holds | Admin, allied medical, domestic, hca, medic, nurse, other, pharmacy, PT/OT, Radiographer (10) | $x_{job}\in\{1, 2, 3, 4, 5, 6,$  $7, 8, 9, 10\}$ |
| Location | Location of individuals job | loc_ed, loc_amu, loc_critcare, loc_elderlycare, loc_infectdis, loc_other, loc_resp_gsm, loc_respiratory (8) | $x_{loc}\in\{1, 2, 3, 4, 5, 6,$  $7, 8\}$ |
| c19zones | Degree of contact with covid 19 persons | “I've not worked in any patient areas”, “I've worked in green zones most days", "I've attended/cared for COVID-19 patients but only after they've been declared no longer infectious”, "I work in red zones most days but don't attend/care for patients myself", "I've attended/cared for patients with COVID-19 in red zones occasionally", "I've attended/cared for patients with COVID-19 in red zones most days" (6) | $x_{zon}\in\{1, 2, 3, 4, 5, 6\}$ |

*Model types*

We consider adjusted and unadjusted versions of three different models (A-C). We considered three separate models instead of performing a Bayesian regression on all primary exposures simultaneously. This is because we found multicollinearity between primary exposures, making parameter inference difficult to interpret when more than one primary exposure was included in the regression analysis model. Unadjusted model A uses only Job as the only covariate function, the adjusted model uses job, age, gender, and ethnicity. Similarly, model B uses zone alone (unadjusted) or together with job, age, and ethnicity (adjusted), and model C looks at location alone (unadjusted) or together with job, age, and ethnicity (adjusted).

*Model parameters*

### Table S4. Summary of the model parameters used in the regression model.

| **Symbol** | **Description** | **Source** | **Value or range** |
| --- | --- | --- | --- |
| *Parameters associated with detection* | | | |
| $n_{se}$ | Number of SARS-CoV-2-positive serological samples tested | Data | 180 |
| $n_{sp}$ | Number of SARS-CoV-2-negative serological samples tested | Data | 650 |
| $y_{se}$ | Number of SARS-CoV-2-positive samples corrected identified as positive | Data | 179 |
| $y_{sp}$ | Number of SARS-CoV-2-negative samples corrected identified as negative | Data | 646 |
| $sp$ | Specificity of serological test | Inferred | 0–1 |
| $se$ | Sensitivity of serological test | Inferred | 0–1 |
| *Parameters associated with seroprevalence* | | | |
| $y_{i}$ | Serostatus of individual *i* | Data | 0 or 1 |
| $\pi_{i, obs}$ | Population-level observed seroprevalence for individual, *i* | Eq. 1 | 0–1 |
| $\pi_{i, true}$ | Population-level true seroprevalence for individual, *i* | Eq. 2 | 0–1 |
| $\beta_{0}$ | Intercept of regression model | Inferred | Real |
| $\beta_{j}^{age}$ | Regression coefficient for the age covariate, category *j*. | Inferred | Real |
| $\beta_{j}^{eth}$ | Regression coefficient for the ethnicity covariate, category *j*. | Inferred | Real |
| $\beta_{j}^{gen}$ | Regression coefficient for the gender covariate, category *j*. | Inferred | Real |
| $\beta_{j}^{job}$ | Regression coefficient for the job covariate, category *j*. | Inferred | Real |
| $\beta_{j}^{zon}$ | Regression coefficient for the zone covariate, category *j*. | Inferred | Real |
| $\beta_{j}^{loc}$ | Regression coefficient for the location covariate, category *j*. | Inferred | Real |

##### Hierarchical model equations

For the standard deviation, we chose exponential distributions with a rate of 1 the prior. This distribution is simple, positive, weakly-informative and has the advantage of weakly-regularising towards zero. The priors for the sensitivity (se) and the specificity (sp) were strongly informed using results from the validation dataset.

Here z is loc, job or zon. Please see **Table S4** for other parameter definitions.

$$(L.1) y_{i} \sim\mathrm{Bernoulli}\left( \pi_{i,obs} \right)$$

$$\left( L.2 \right) y_{se} \sim\mathrm{Bin}\left( n_{se}, se \right)$$

$$(L.3) y_{sp} \sim\mathrm{Bin}\left( n_{sp}, sp \right)$$

$${(Eq.1) \pi}_{i, obs}= {se*\pi}_{i,true}+\left( 1-\pi_{i,true} \right)*\left( 1-sp \right)$$

$$\left( Eq. 2 \right)\mathrm{logit}\left( \pi_{i,true} \right)=\beta_{0}+\beta_{x_{i,z}}^{z}$$

$${(Pr. 1) \beta}_{0}\sim N\left( 0,1.5 \right)$$

$$\beta_{j}^{z}\sim N\left( 0,\sigma_{z} \right)$$

$$\sigma_{z}\sim\mathrm{Exponential}(1)$$

$$se \sim N(5, 1)$$

$$sp \sim N(5, 1)$$

*Adjusted*

$$(L.1) y_{i} \sim\mathrm{Bernoulli}\left( \pi_{i,obs} \right)$$

$$\left( L.2 \right) y_{se} \sim\mathrm{Bin}\left( n_{se}, se \right)$$

$$(L.3) y_{sp} \sim\mathrm{Bin}\left( n_{sp}, sp \right)$$

$${(Eq.1) \pi}_{i, obs}= {se*\pi}_{i,true}+\left( 1-\pi_{i,true} \right)*\left( 1-sp \right)$$

$$\left( Eq. 2 \right)\mathrm{logit}\left( \pi_{i,true} \right)=\beta_{0}+\beta_{x_{i,age}}^{age}+\beta_{x_{i,eth}}^{eth}+\beta_{x_{i,gen}}^{gen}+\beta_{x_{i,z}}^{z}$$

$$\beta_{0}\sim N\left( 0,1.5 \right)$$

$$\beta_{j}^{age}\sim N\left( 0,\sigma_{age} \right)$$

$$\beta_{j}^{eth}\sim N\left( 0,\sigma_{eth} \right)$$

$$\beta_{j}^{gen}\sim N\left( 0,\sigma_{gen} \right)$$

$$\beta_{j}^{z}\sim N\left( 0,\sigma_{z} \right)$$

$$\sigma_{age}\sim\mathrm{Exponential}(1)$$

$$\sigma_{eth}\sim\mathrm{Exponential}(1)$$

$$\sigma_{gen}\sim\mathrm{Exponential}(1)$$

$$\sigma_{z}\sim\mathrm{Exponential}(1)$$

$$se \sim N(5, 1)$$

$$sp \sim N(5, 1)$$

##### Implementation and model fit

Using stan programme language and the cmdstanr interface, we obtained posterior distributions by implementing a Hamiltonian Monte Carlo algorithm four chains with 2,000 iterations each (1,000 burn in). We implemented the code using a non-centralised parameterisation to prevent divergent chains. After numerous simulations for the three different submodels, adjusted and unadjusted, between 0/4000 and 5/4000 (0%) chains ended with divergence, indicating the Hamiltonian Monte Carlo methods behaved well. The posterior distributions shown in this paper and on the Github repository are taken from a simulation with 0 divergent chains. Further the Rhat values for all of the parameters were less than 1.1, implying the four chains have mixed well.

##### Marginalisation

In order to determine the true seroprevalence for a single covariate in the adjusted models, we integrate out the other covariates. For example, to get the marginal posterior for the seroprevalence of age group j we calculate,

$${\pi_{j,age}= \int_{1}^{4} \int_{1}^{2} \int_{1}^{z} {(\beta}_{0}+\beta_{x_{1}}^{z}+\beta_{x_{2}}^{gen}+\beta_{x_{3}}^{eth}+\beta_{j}^{age}){dx}_{1}dx_{2}dx_{3}}$$

Using the posterior distributions for $\beta_{x_{i}}^{j}$,. In practice this can be calculated through

$$\pi_{j,age}=\sum_{x_{3}=1}^{4} \sum_{x_{2}=1}^{2} \sum_{x_{1}=1}^{8} {(\beta}_{0}+\beta_{x_{1}}^{z}+\beta_{x_{2}}^{gen}+\beta_{x_{3}}^{eth}+\beta_{j}^{age})p_{gen}\left[ x \right]p_{eth}\left[ x \right]p_{z}\left[ x \right]$$

Where $p_{z}=(p_{z}\left[ 1 \right], p_{z}\left[ 2 \right], \ldots{,p}_{z}\left[ n \right])$ is a simplex vector of the proportions of individuals in each group of covariate z.

#### Seroprevalence model B — Symptomatic

##### Model outline

A similar model was used to estimate the proportion of asymptomatic infections. In the regression part the model remains the same but the prevalence is no longer adjusted according to sensitivity and specificity, and the response is the binary outcome 0 = asymptomatic and 1 symptomatic. The three models (A-C) are adjusted and unadjusted as before. The model equations and diagram are given below.

*Data*

We included all samples which were seropositive at their first bleed and completed the study (n = 246).

*Model variables*

The response variable is the symptomatic status of an individual, with 0 = asymptomatic, and 1 = symptomatic. The covariate variables remain the same.

*Model types*

Same as Prevalence model A .

##### Hierarchical model

*Unadjusted*

Where $j_{k}$ are the indicator functions for the respective covariate groups:

$$(L.1) y_{i} \sim\mathrm{Bernoulli}\left( \pi_{i,obs} \right)$$

$$\left( Eq. 2 \right)\mathrm{logit}\left( \pi_{i,true} \right)=\beta_{0}+\beta_{z\left[ j_{4} \right]}x_{i, z}$$

$${(Pr. 1) \beta}_{0}\sim N\left( 0,1.5 \right)$$

$$\beta_{j}^{z}\sim N\left( 0,\sigma_{z} \right)$$

$$\sigma_{z}\sim\mathrm{exponential}(1)$$

*Adjusted*

$$(L.1) y_{i} \sim\mathrm{Bernoulli}\left( \pi_{i,obs} \right)$$

$$\left( Eq. 2 \right)\mathrm{logit}\left( \pi_{i,true} \right)=\beta_{0}+\beta_{x_{i,age}}^{age}+\beta_{x_{i,eth}}^{eth}+\beta_{x_{i,gen}}^{gen}+\beta_{x_{i,z}}^{z}$$

$${(Pr. 1) \beta}_{0}\sim N\left( 0,1.5 \right)$$

$$\beta_{x_{i,age}}^{age}\sim N\left( 0,\sigma_{age} \right)$$

$$\beta_{x_{i,eth}}^{eth}\sim N\left( 0,\sigma_{eth} \right)$$

$$\beta_{x_{i,gen}}^{gen}\sim N\left( 0,\sigma_{gen} \right)$$

$$\beta_{x_{i,z}}^{z}\sim N\left( 0,\sigma_{z} \right)$$

$$\sigma_{age}\sim\mathrm{Exponential}(1)$$

$$\sigma_{eth}\sim\mathrm{Exponential}(1)$$

$$\sigma_{gen}\sim\mathrm{Exponential}(1)$$

$$\sigma_{z}\sim\mathrm{Exponential}(1)$$

##### Implementation and model fit

Same as Prevalence model A.

##### Marginalisation

Same as Prevalence model A.

#### Antibody kinetics model

##### Model outline

*Data*

We included all samples which included a positive serological sample at their first and second bleed (n = 264). We consider four different antibody-antigen interactions , spike-IgG, NCP-IgG, spike-IgA, NCP-IgA. For the IgG, the units are in universal WHO-recommended units, for the IgA the units are standardised according to the values in the study.

*Model variables*

### Table S5. Summary of the response variables and the covariates used in the regression model.

The elements in the encoding vector are associated with the groups given in the categories column in the order they are written.

| **Variable** | **Description** | **Values** | **Symbol** |
| --- | --- | --- | --- |
| *Response* | | | |
| Antibody units at first bleed for measure *m* | Log_2_ of the universal unit titres at first bleed. | 0–9.966* | $y_{s}^{m}$ |
| Change in antibody levels between bleeds for measure *m* | Difference in log of the universal units titres | -9.966 and 9.966 | $y_{d}^{m}$ |
| *Covariates* | | | |
| Age | Age category of individual | <30, 30-39, 40-49, 50-59, 60+ years (5) | $x_{age}\in\{1, 2, 3, 4, 5\}$ |
| Ethnicity | Ethnicity of individual | White, Black, Asian, Other (4) | $x_{eth}\in\{1, 2, 3, 4\}$ |
| Gender | Gender of individual | Female , Male (2) | $x_{gen}\in\{1, 2\}$ |
| Disease severity | Whether to individual experienced symptoms | Asymptomatic, symptomatic (2) | $x_{sym}\in\{1, 2\}$ |

*Model types*

We consider two different regression models, one using the antibody units at the first bleed, and another using the change in antibody levels between bleeds. For each of these models, the regression is performed for four different response variables, one for each antibody-antigen interaction. The covariates are age, ethnicity, gender and disease severity.

*Model parameters*

### Table S6. Summary of the model parameters used in the regression model.

| **Symbol** | **Description** | **Source** | **Value** |
| --- | --- | --- | --- |
| *Parameters associated with seroprevalence* | | | |
| $y_{s, i}^{m}$ | Log antibody units at first bleed of individual i, for measure m | Data | 0–9.966 |
| $y_{d, i}^{m}$ | Log antibody units at first bleed of individual i, for measure m | Data | -9.966–9.966 |
| T | Time between bleeds in days. | Data | Integer |
| $\beta_{0}$ | Intercept of regression model | Inferred | Real |
| $\beta_{j}^{age}$ | Regression coefficient for the age covariate, category *j*. | Inferred | Real |
| $\beta_{j}^{eth}$ | Regression coefficient for the ethnicity covariate, category *j*. | Inferred | Real |
| $\beta_{j}^{gen}$ | Regression coefficient for the gender covariate, category *j*. | Inferred | Real |
| $\beta_{j}^{sym}$ | Regression coefficient for the symptomatic covariate, category *j*. | Inferred | Real |
| $\sigma_{a}$ | Standard deviation between age groups. | Inferred | >0 |
| $\sigma_{e}$ | Standard deviation between ethnic groups. | Inferred | >0 |
| $\sigma_{g}$ | Standard deviation between gender groups. | Inferred | >0 |
| $\sigma_{s}$ | Standard deviation between symptomatic groups. | Inferred | >0 |

##### Hierarchical model equations

*Antibodies at first bleed*

$$x_{i} \sim N(\mu_{i}, \sigma)$$

$$\mu_{i}= \beta_{0}+\beta_{x_{i,age}}^{age}+\beta_{x_{i,eth}}^{eth}+\beta_{x_{i,gen}}^{gen}+\beta_{x_{i,sym}}^{sym}$$

$$\beta_{0}\sim N(0.0, 0.2)$$

$$\beta_{x_{i,age}}^{age}\sim N\left( 0, \sigma_{a} \right)$$

$$\beta_{x_{i,eth}}^{eth} \sim N\left( 0, \sigma_{e} \right)$$

$$\beta_{x_{i,gen}}^{gen} \sim N\left( 0, \sigma_{g} \right)$$

$$\beta_{x_{i,sym}}^{sym} \sim N\left( 0, \sigma_{s} \right)$$

$$\sigma_{a} \sim Exponential(1)$$

$$\sigma_{e} \sim Exponential(1)$$

$$\sigma_{g} \sim Exponential(1)$$

$$\sigma_{s} \sim Exponential(1)$$

*Difference at between bleeds*

$$x_{i} \sim N(\mu_{i}, \sigma)$$

$$\mu_{i}= T*(\beta_{0}+\beta_{x_{i,age}}^{age}+\beta_{x_{i,eth}}^{eth}+\beta_{x_{i,gen}}^{gen}+\beta_{x_{i,sym}}^{sym})$$

$$\beta_{0}\sim N(0.0, 0.2)$$

$$\beta_{x_{i,age}}^{age}\sim N\left( 0, \sigma_{a} \right)$$

$$\beta_{x_{i,eth}}^{eth} \sim N\left( 0, \sigma_{e} \right)$$

$$\beta_{x_{i,gen}}^{gen} \sim N\left( 0, \sigma_{g} \right)$$

$$\beta_{x_{i,sym}}^{sym} \sim N\left( 0, \sigma_{s} \right)$$

$$\sigma_{a} \sim Exponential(1)$$

$$\sigma_{e} \sim Exponential(1)$$

$$\sigma_{g} \sim Exponential(1)$$

$$\sigma_{s} \sim Exponential(1)$$

##### Implementation

Same as Prevalence model A.

##### Marginalisation

Same as Prevalence model A

#### Heterogenous sensitivity model

##### Model outline

In this model we use a binomial model with a gaussian process prior to model the relationship between the sensitivity/specificity of the assay validation dataset and the implied sensitivity/specificity of the HERO dataset.

*Data*

This model uses the same validation dataset as the IgG ELISA, which comprises 865 samples for which the serostatus of the individual is known (190 positive and 675 negative), and associated covariate data includes *A*_450_ value

*Model variables*

### Table S7. Summary of the model parameters used in the Heterogenous sensitivity model.

| **Variable** | **Values** | **Symbol** |
| --- | --- | --- |
| *Variables associated with control dataset* | | |
| *A*_450_ value of sample number i*,* from known SARS-CoV-2 positive individual against protein *p* in the assay validation dataset. | 1.8–4.5 | x^+,p^_VD,i_ |
| *A*_450_ value of sample number *i* from known SARS-CoV-2 negative individual against protein *p* in the assay validation dataset. | 0–1.8 | x^-,p^_VD,i_ |
| Number of samples from known SARS-CoV-2 positive individuals in the assay validation dataset. | 151 | N^+,p^_VD_ |
| Number of samples from known SARS-CoV-2 negative individuals in the assay validation dataset. | 160 | N^-,p^_VD_ |
| Sensitivity of the assay validation dataset against protein *p*. | {60, 61, …, 100} | X^p^ _jse_ |
| Specificity of the assay validation dataset against protein *p*. | {60, 61, …, 100} | X^p^_jsp_ |
| *Variables associated with the HERO dataset* | | |
| *A*_450_ value of sample number *i* from individuals in the HERO dataset who are predicted to be SARS-CoV-2 positive against protein *p* from the HERO ELISA. | 1.8–4.5 | x^+^_HERO, i_ |
| *A*_450_ value of sample number *i* from individuals in the HERO dataset who are predicted to be SARS-CoV-2 negative against protein *p* from the HERO ELISA. | 0–1.8 | x^+^_HERO, i_ |
| Number of samples from individuals in the assay validation dataset who are predicted to be SARS-CoV-2 positive from the HERO ELISA. | 311 | N^+^_HERO_ |
| Number of samples from individuals in the assay validation dataset who are predicted to be SARS-CoV-2 negative from the HERO ELISA. | 964 | N^-^_HERO_ |
| Implied sensitivity of the HERO data set | 0–100 | Y^p^_jse_ |
| Implied sensitivity of the HERO data set | 0–100 | Y^p^_jsp_ |
| Age-dependent v*ariables associated with the HERO dataset* | | |
| Number of samples from individuals in the assay validation dataset who are predicted to be SARS-CoV-2 positive from the HERO ELISA, in age group *a*. | {67, 69, 72, 76, 27} | N^+, a^_HERO_ |
| Number of samples from individuals in the assay validation dataset who are predicted to be SARS-CoV-2 negative from the HERO ELISA, in age group *a*. | {200, 237, 242, 238, 47} | N^-, a^_HERO_ |
| Implied sensitivity of the HERO data set, in age group *a*. | 0–100 | Y^p, a^_jse_ |
| Implied sensitivity of the HERO data set, in age group *a*. | 0–100 | Y^p, a^_jsp_ |

##### Model equations

The value for the implied sensitivity and specificity for an antigen protein *p* given by

$Y_{j_{se}}^{p}=\frac{\sum_{i=1}^{N_{HERO}^{+}} \boldsymbol{1}_{\boldsymbol{\{}x_{HERO,i}^{+}\boldsymbol{>}T_{j_{se}}^{p}\boldsymbol{\}}}}{N_{HERO}^{+}}$, where $A_{j_{se}}^{p}$, the absorption value associated with a sensitivity of *j*, is given by $\{A_{j_{se}}^{p}: \frac{\sum_{i=1}^{N_{VD}^{+}} \boldsymbol{1}_{\left( x_{VD,i}^{+,p}\boldsymbol{>}A_{j_{se}}^{p} \right)}}{N_{VD}^{+}}=X_{j_{se}}^{p}\}$

$Y_{j_{sp}}^{p}=\frac{\sum_{i=1}^{N_{HERO}^{+}} \boldsymbol{1}_{\boldsymbol{\{}x_{HERO,i}^{+}\boldsymbol{<}A_{j_{sp}}^{p}\boldsymbol{\}}}}{N_{HERO}^{+}}$, where $A_{j_{sp}}^{p}$, the absorption value associated with a specificity of *j*, is given by $\{A_{j_{sp}}^{p}: \frac{\sum_{i=1}^{N_{VD}^{+}} \boldsymbol{1}_{\left( x_{VD,i}^{+,p}\boldsymbol{<}A_{j_{sp}}^{p} \right)}}{N_{VD}^{+}}=X_{j_{sp}}^{p}\}$

To model the implied sensitivity and specificity we use a Gaussian process model with a multivariate normal realisation:

$$\mathrm{logit}\left( p_{j_{se}}^{p} \right)= f_{j_{se}}^{p} \sim MVN(\boldsymbol{0}, \boldsymbol{K}\left( X_{j_{se}}^{p} | \alpha^{p},\rho^{p} \right))$$

$$Y_{j_{se}}^{p} \sim Bin(N_{HERO}^{+}, p_{j_{se}}^{p})$$

Here $\boldsymbol{K}\left( X_{se, i} \right|\alpha^{p},\rho^{p})$ is an exponential quadratic covariance function with a small positive constant added to the diagonal to ensure positive definiteness,

$${\boldsymbol{K}({x \left| \alpha^{p},\rho^{p} \right)}_{i,j}= {(\alpha}^{p})}^{2}\exp\left( -\frac{1}{2{{(\rho}^{p})}^{2}}(X_{i}^{p}-X_{j}^{p}) \right)+ \delta_{i, j}\sigma^{2}$$

with priors given by

$$\rho^{p} \sim\mathrm{InvGamma}(5, 5)$$

$$\alpha^{p} \sim N(0, 1)$$

for an antigen protein *p*.

*Age-dependent model*

In the age dependent model, we consider the age category associated with each samples in the HERO dataset, *a*, and derive the value of $Y_{j_{se}}^{p,a}$ , $Y_{j_{sp}}^{p,a}$ from

$Y_{j_{se}}^{p,a}=\frac{\sum_{i=1}^{N_{HERO}^{+, a}} \boldsymbol{1}_{\boldsymbol{\{}x_{HERO,i}^{+, a}\boldsymbol{>}{OD}_{j_{se}}^{p}\boldsymbol{\}}}}{N_{HERO}^{+, a}}$, $Y_{j_{sp}}^{p,a}=\frac{\sum_{i=1}^{N_{HERO}^{-, a}} \boldsymbol{1}_{\boldsymbol{\{}x_{HERO,i}^{-, a}\boldsymbol{<}{OD}_{j_{sp}}^{p}\boldsymbol{\}}}}{N_{HERO}^{-, a}}$, where $A_{j_{se}}^{p}$is as given above.

In the age-dependent model a multioutput gaussian process prior is used which is realised by a 40 x 5 dimensional matrix normal distribution:

$$\mathrm{logit}\left( p_{j_{se}}^{p, a} \right)= f_{j_{se}}^{p, a} \sim MatrixNormal(\boldsymbol{0}, \boldsymbol{K}\left( X_{j_{se}}^{p} | \alpha^{p},\rho^{p} \right), C(\phi))$$

$$Y_{j_{se}}^{p,a} \sim Bin(N_{HERO}^{+}, p_{j_{se}}^{p, a})$$

In which the prior for $C\left( \phi\right)$is LKJCorr(3). Note that $f_{j_{se}}^{p, a}\in\mathbb{R}^{40 \times5}$, where 40 is the number of dimensions of the sensitivity/specificity and 5 is the number of age groups.

##### Implementation and model fit

Same as Prevalence model A.

### Figure S3. Study flow diagram

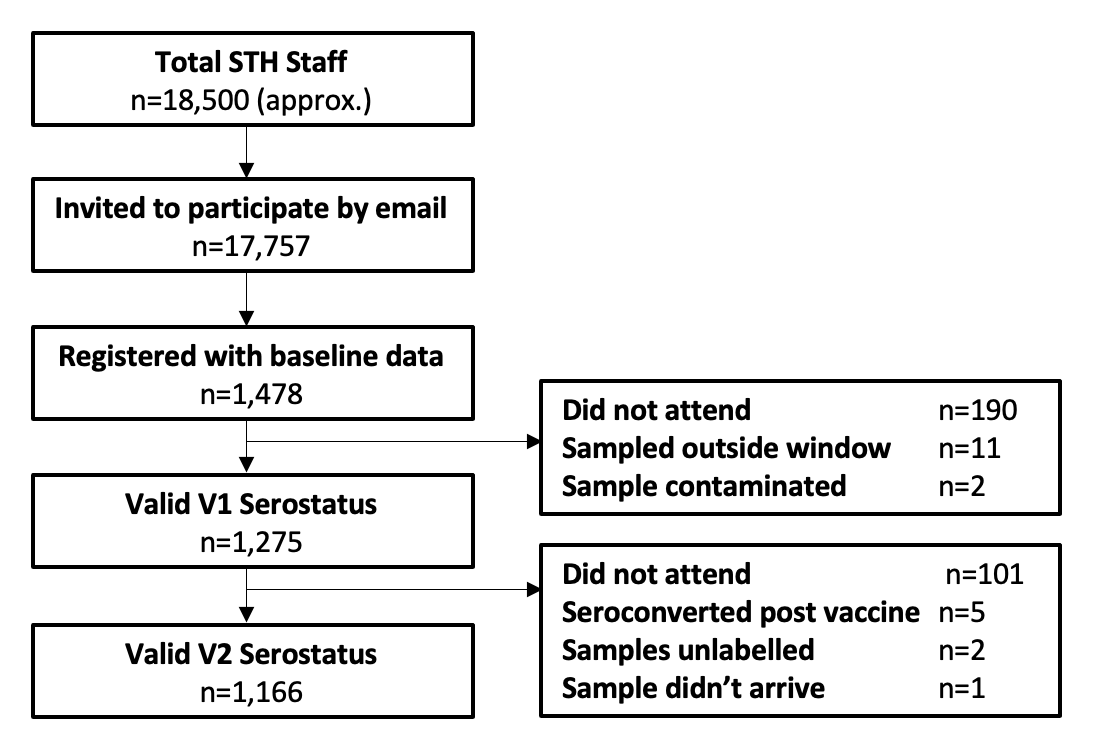

### Figure S4. Histogram (overlayed) showing the symptom onset, date of first bleed (all cases and symptomatic cases only), and time at second bleed (all cases and symptomatic cases only).

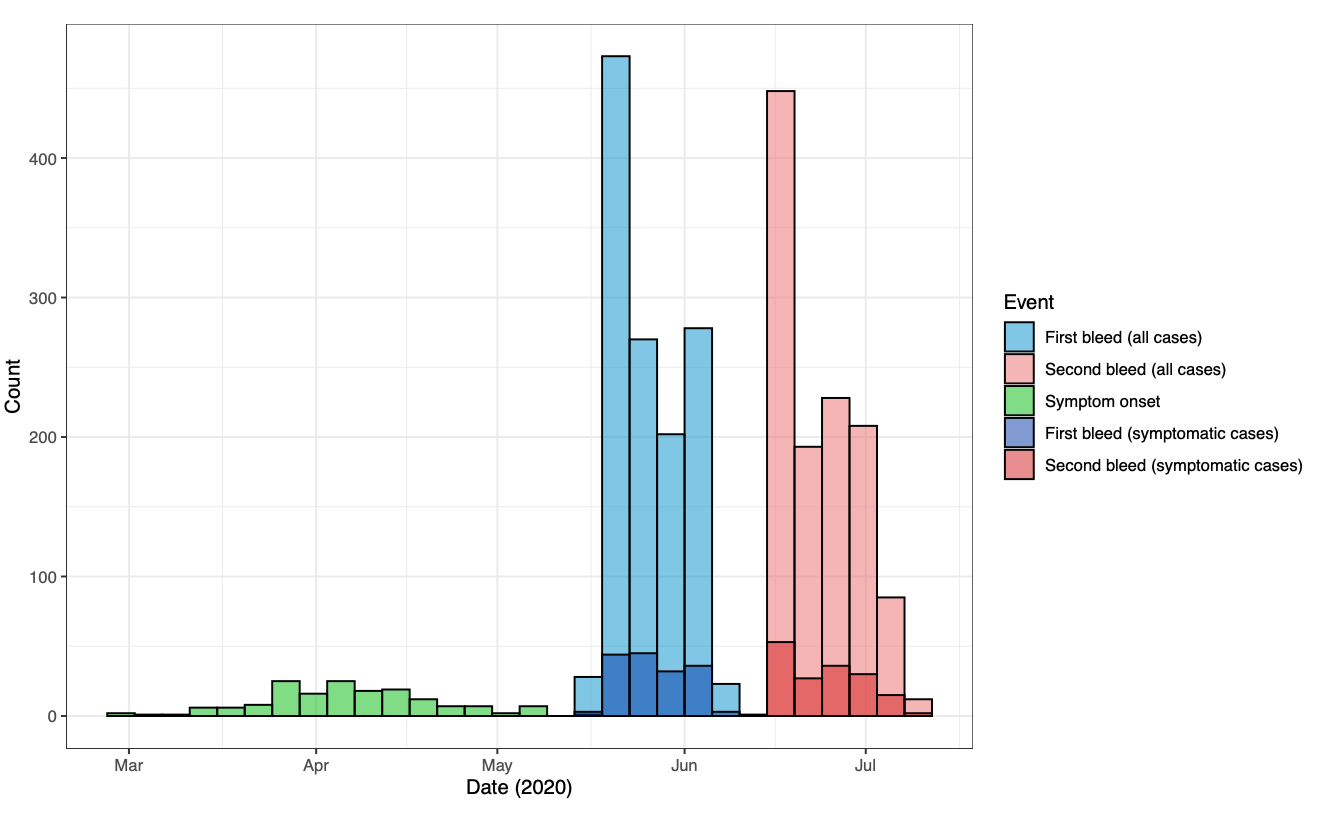

### Figure S5. Model-predicted proportion of asymptomatic estimates for three different models (A-C), adjusted and unadjusted with covariates gender, age group and ethnicity.

Black stars represent point values from the data. The point and whiskers represent the mean value and 95% CrI of the posterior distribution. The three models differed by their primary exposure, Model A used covid-19 zones, Model B the job role, and Model C the job location. Each model was evaluated either unadjusted (primary exposure only) or adjusted (primary exposure with age, gender, and ethnicity).

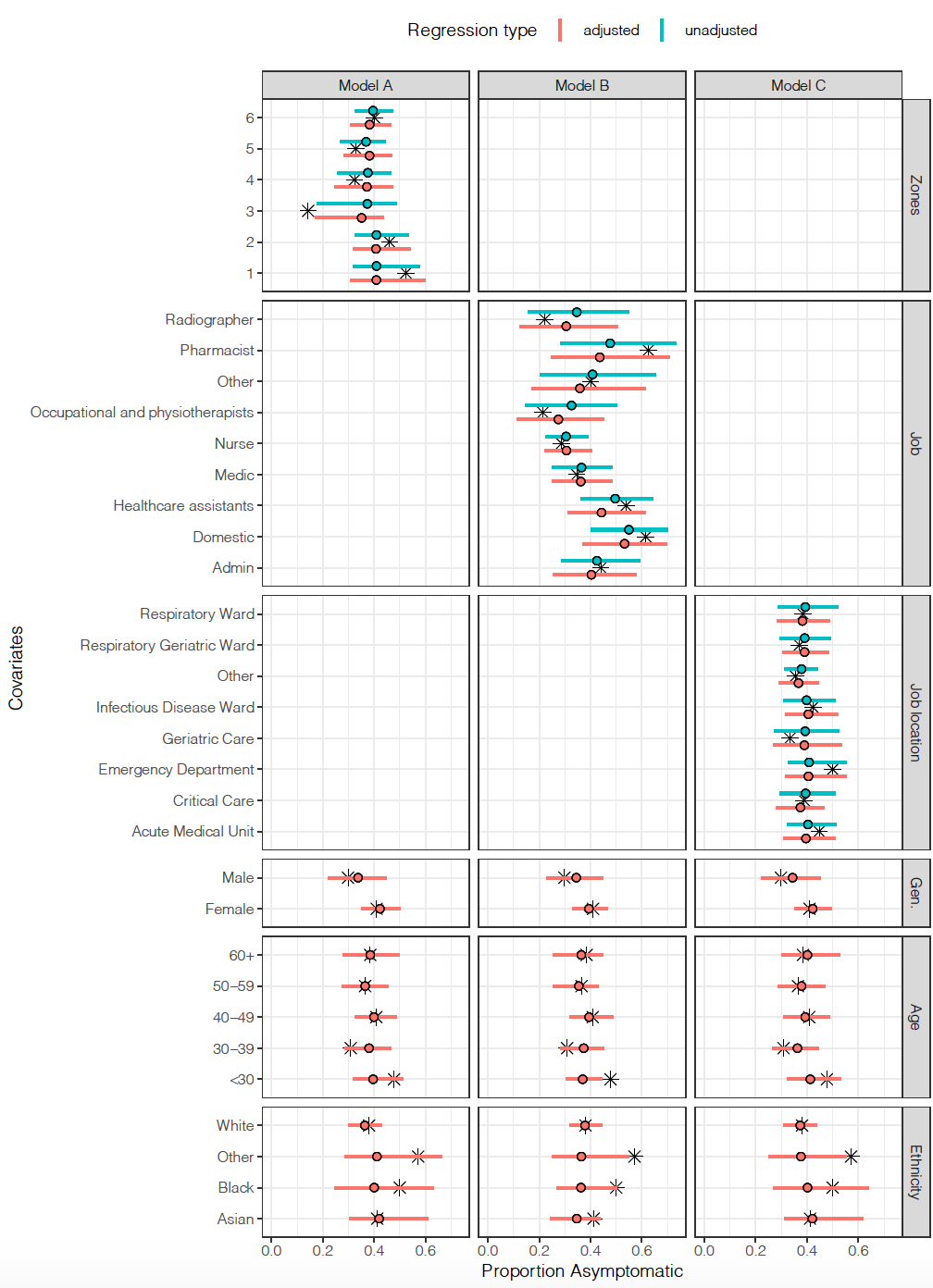

### Figure S6. Correlation between the four different antibody measures for 264 serological samples.

Measures have all been standardised such that the mean is 0 and standard deviation is 1. Labeled values on each facet shows the R-squared values.

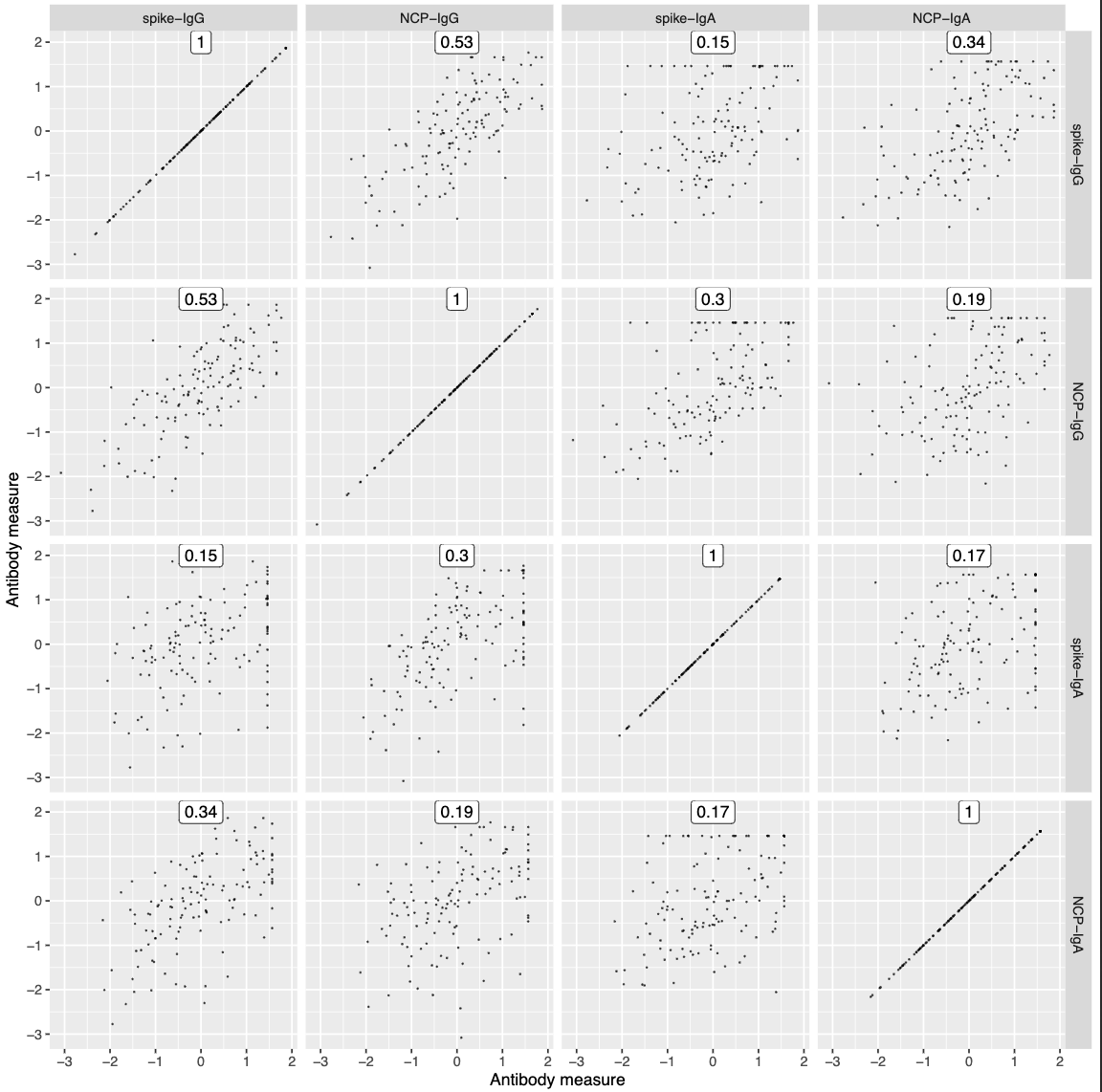

### Figure S7. Rate of decline for the antibody concentrations post-symptom onset for the four antibody measures. The fitted line is from a linear regression, with the 95% CI shown in red.

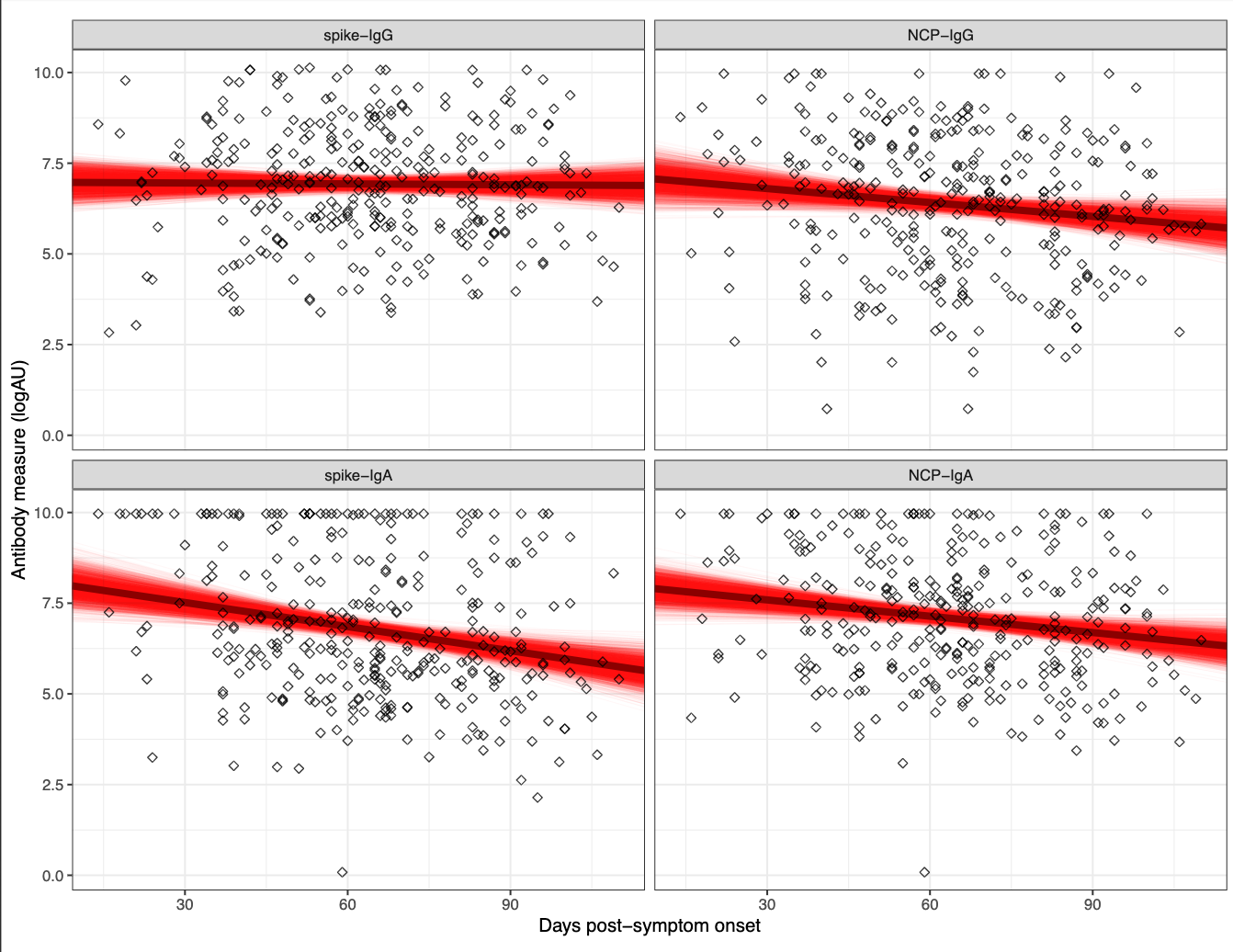

### Figure S8. Relationship between sensitivity/specificity and the cutoff value for the control dataset.

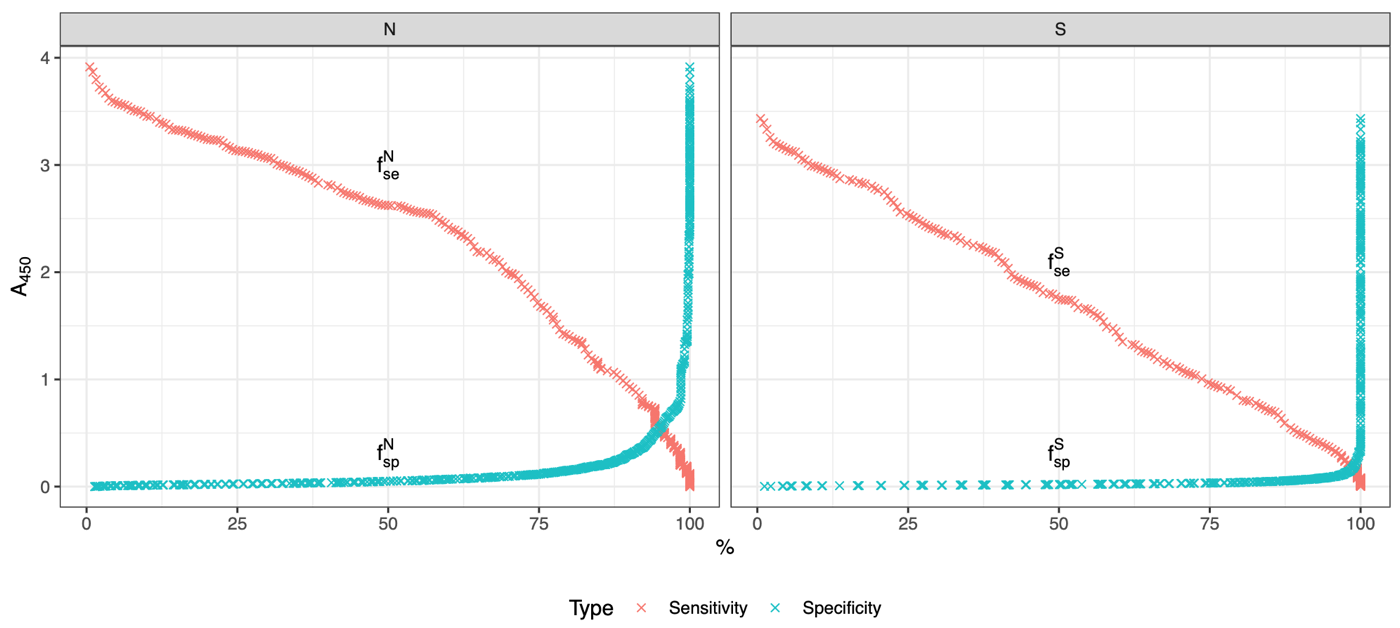

### Figure S9. ROC curves with the *A*_450_ cut-off value indicated in red for the control dataset. x-axis shows the False Positive Rate, y-axis is the sensitivity.

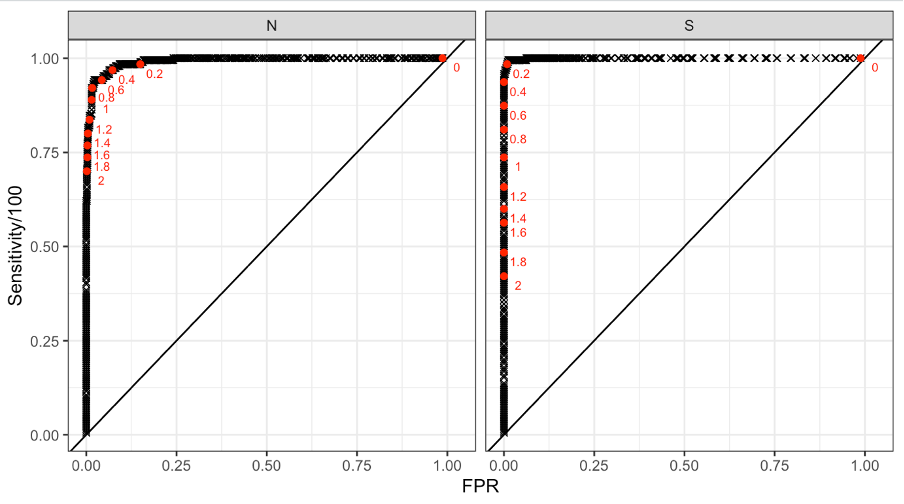

### Figure S10. ROC curves for different age groups and antigen proteins, with the *A*_450_ cut-off value indicated in various colours for the control dataset.

x-axis shows the False Positive Rate, y-axis is the Sensitivity.
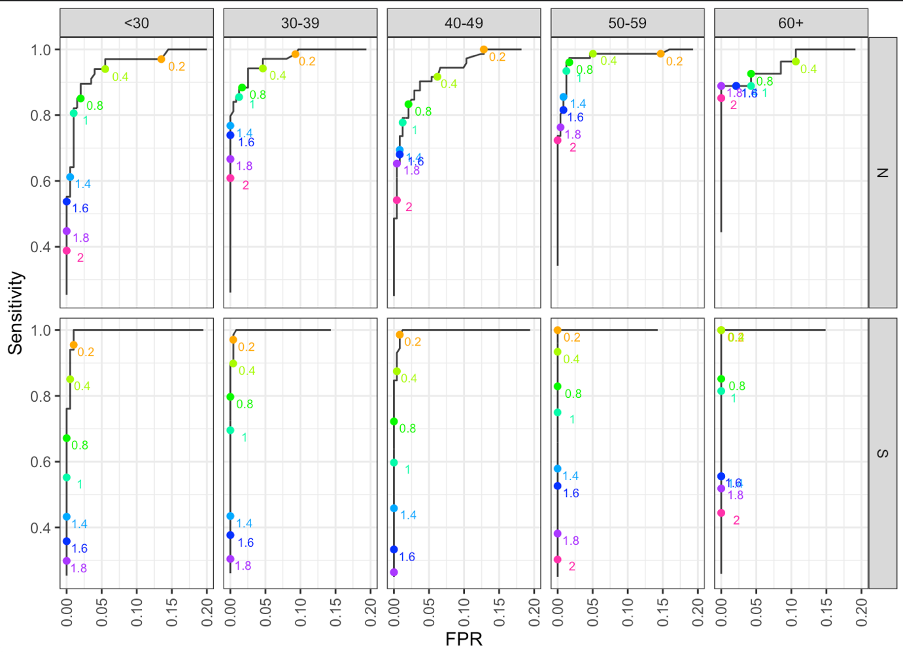

### Figure S11. (a) Specificity of the control data set against the implied specificity of the HERO dataset for spike and nucleoprotein. (b) Specificity of the control data set against the implied age-specific specificity of the HERO dataset for spike and nucleoprotein.

Black line and ribbon shows median and 95% CrI for the posterior distributions respectively.
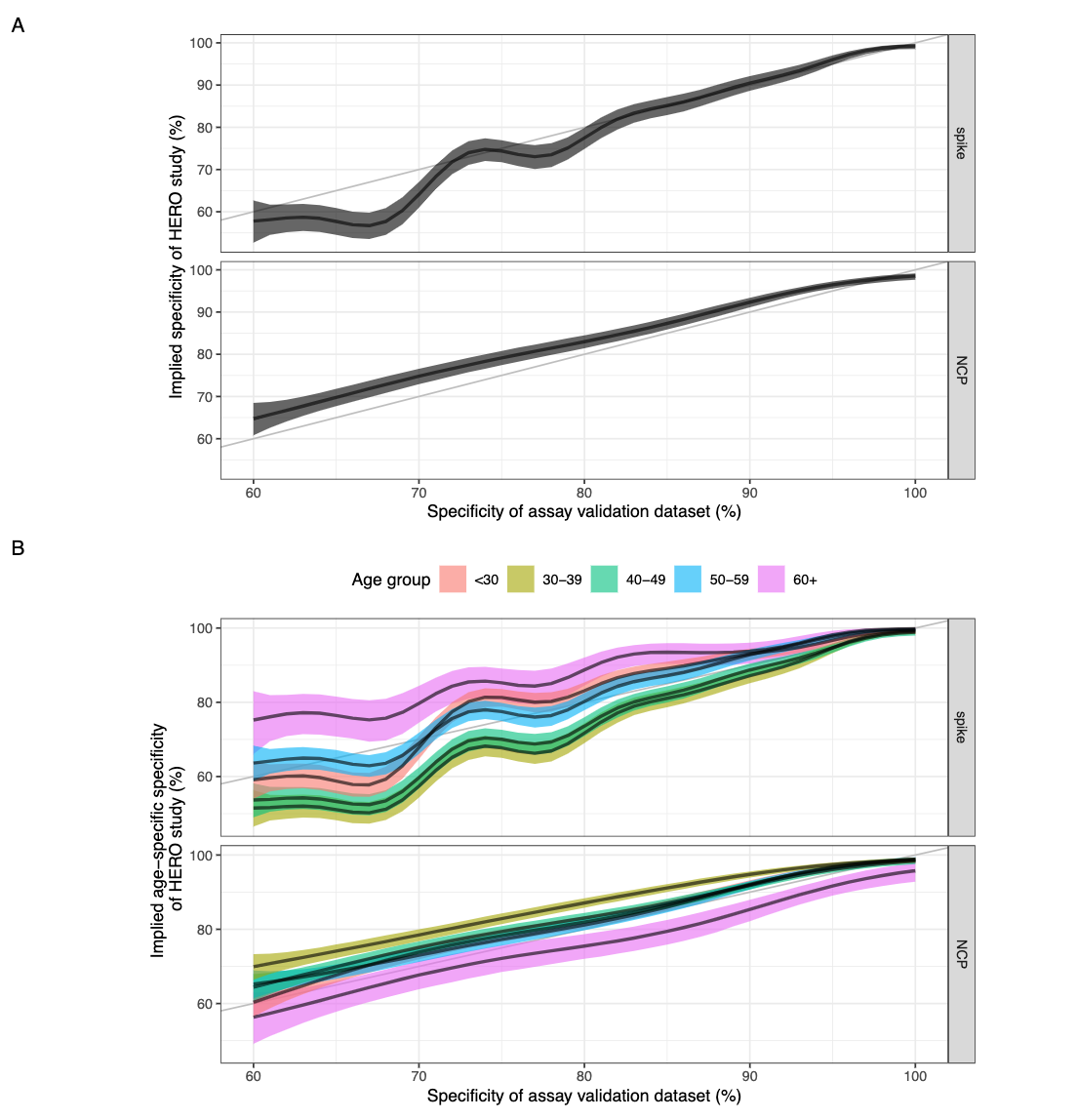

#### References

1. Hanson, K.E., et al., *Infectious Diseases Society of America Guidelines on the Diagnosis of COVID-19: Serologic Testing.* Clin Infect Dis, 2020.

2. Mattiuzzo et al*., Establishment of the WHO International Standard and Reference Panel for anti-SARS-CoV-2 antibody.* 2020, WHO Expert Committee on Biological Standardization. WHO/BS/2020.2403

3.	Tugaeva KV, Hawkins DEDP, Smith JLR, Bayfield OW, Ker DS, Sysoev AA, Klychnikov OI, Antson AA, Sluchanko NN. The Mechanism of SARS-CoV-2 Nucleocapsid Protein Recognition by the Human 14-3-3 Proteins. *J Mol Biol*. 2021 433, 166875. doi: 10.1016/j.jmb.2021.166875.

4.	Studier, FW (2018) in *Current Protocols in Molecular Biology*, e63. doi: 10.1002/cpmb.63

5.	Sayers, JR Viral polymerase-associated 5′–3′ exonucleases: expression, purification, and uses. *Methods in Enzymol.* 275, 227–238 (1996)
